# Mechanism of optimal time-course COVID-19 vaccine prioritization based on non-Markovian steady-state prediction

**DOI:** 10.1101/2021.10.11.21264836

**Authors:** Mi Feng, Liang Tian, Changsong Zhou

## Abstract

Vaccination is essential for controlling the coronavirus disease (COVID-19) pandemic. An effective time-course strategy for the allocation of COVID-19 vaccines is crucial given that the global vaccine supply will still be limited in some countries/regions in the near future and that mutant strains have emerged and will continue to spread worldwide. Both asymptomatic and symptomatic transmission have played major roles in the COVID-19 pandemic, which can only be properly described as a typical non-Markovian process. However, the prioritization of vaccines in the non-Markovian framework still lacks sufficient research, and the underlying mechanism of the time-course vaccine allocation optimization has not yet been uncovered. In this paper, based on an age-stratified compartmental model calibrated through clinical and epidemiological data, we propose optimal vaccination strategies (OVS) through steady-state prediction in the non-Markovian framework. This OVS outperforms other empirical vaccine prioritization approaches in minimizing cumulative infections, cumulative deaths, or years of life lost caused by the pandemic. We found that there exists a fast decline in the prevention efficiency of vaccination if vaccines are solely administered to a selected age group, which indicates that the widely adopted strategy to continuously vaccinate high-risk group is not optimal. Through mathematical analysis of the model, we reveal that dynamic vaccine allocations to combinations of different age groups is necessary to achieve optimal vaccine prioritization. Our work not only provides meaningful references for vaccination in countries currently lacking vaccines and for vaccine allocation strategies to prevent mutant strains in the future, but also reveals the mechanism of dynamic vaccine allocation optimization, forming a theoretical and modelling framework empirically applicable to the optimal time-course prioritization.

## Introduction

Coronavirus disease (COVID-19), caused by severe acute respiratory syndrome coronavirus 2 (SARS-CoV-2), has spread to more than 200 countries/regions worldwide, with up to 231 million confirmed cases and 4.75 million deaths as of 26 September 2021 [1]. While different countries/regions have enacted different containment and mitigation strategies, including school closures, travel bans, and house quarantine, vaccination is the ultimate strategy to reduce the disease incidence and resume normal socio-economic activities. As of early October 2021, approximately 6.3 billion doses of COVID-19 vaccines have been administered worldwide, and approximately 44.5% of the global population has received at least one dose of vaccine, but only 2.3% of people in low-income countries/regions have received at least one dose [2].

Different countries/regions implement different vaccination policies according to the situation of virus spread and availability of vaccines. However, vaccine shortages in some countries/regions remain a major issue in the current phase, despite scaling up vaccine production. As a result, it remains important to create effective strategies for vaccine distribution to minimize the burden of COVID-19 under different situations of disease spread and availability of vaccines [3–12]. Research in this line is also of long-term impact for future pandemic. Vaccination is performed according to two major approaches: direct protection, in which individuals with a high risk of disease and death are vaccinated first; and indirect protection, which prioritizes those with occupations associated with high transmission [3]. Bubar *et al*. evaluated the impact of five COVID-19 vaccine prioritization strategies and found that vaccinating adults aged 20–49 years is the best approach to reduce the cumulative incidence of infection, while prioritizing vaccines to those > 60 years old can minimize the number of deaths and years of life lost [3]. Matrajt *et al*. showed that direct vaccination to high-risk age groups can achieve optimal results in terms of reducing deaths when the vaccine effectiveness is low, regardless of vaccination coverage; however, they also highlighted the importance of distributing vaccines to high-transmission age groups for higher vaccine effectiveness and vaccination coverage [4]. Buckner *et al*. illustrated that younger essential workers should be prioritized to prevent the spread of disease, while older people need to be vaccinated first to control the number of deaths [5]. The vaccination strategy in the United States in early stage was a combination of direct protection and indirect protection, meaning that vaccines are prioritized for those in high-risk age groups and those with high-transmission-risk occupations simultaneously [13]. However, it remains unclear if the empirical strategies are most effective and what the optimal strategies are. Searching for optimal strategies is not feasible without a model that is well-grounded in real data.

Previous studies on vaccination prioritization for preventing COVID-19 are mostly limited to the memoryless Markovian modelling framework, where the dynamics process that individuals contract a virus, recover, and die of the disease are regarded as Poisson processes characterized by exponential distributions of event time. However, the current COVID-19 pandemic possesses strong non-Markovian characteristics in that there is asymptomatic transmission and the infectiousness of an infected individual varies over time from the point of being infected [14–16]. These strong non-Markovian characteristics can significantly affect the dynamics of disease spread in contrast to the Markovian dynamics [17–22]. Therefore, to overcome the limitations of Markovian spreading dynamics on the assessment of vaccination strategy, our analysis places emphasis on age-stratified vaccination optimization in data-calibrated non-Markovian spreading dynamics. In addition, although Buckner *et al*. [5] attempted to search for dynamic optimal vaccine allocations to impede virus spread by using optimization algorithms, the underlying mechanism of the dynamic optimal vaccine allocation has not been analysed. Therefore, it remains unknown why the optimal vaccine allocation is dynamic, what prompts the vaccination of age group combinations, and how the previous vaccination optimization will affect future optimization. Uncovering the evolution mechanism of optimal strategies will deepen our understanding of time-course vaccine prioritization and provide more fundamental guidance for further prevention of disease.

Our model is calibrated against the clinical characteristics of COVID-19 progression and transmission, real contact data between different age groups, as well as the vaccine efficacy and realistic continuous implementation of vaccinations [2,23]. The data-calibrated non-Markovian model allows all infection dynamics in asymptomatic, symptomatic, and quarantine periods to be integrated into a single infection process, so as to treat the infected individuals as one state. Meanwhile, the recovery and death processes in our model are described as one removal process. The non-Markovian model imposes many challenges in searching for an optimal vaccine distribution strategy, mainly because of its insolvability, which can inhibit the theoretical vaccine allocation optimization. Here, we develop a method to accurately predict the steady state without calculating the total transient process, which enables us to overcome the challenge of non-Markovian optimization. We theoretically determine the globally dynamic optimal vaccine allocation strategies to respectively minimize different metrics, i.e., cumulative infections (CI), cumulative deaths (CD), or years of life lost (YLL), at steady state, and compare them with several other empirical strategies. Theoretical analysis of the dynamic decline in the vaccination efficiency and the shift of the priority groups reveals the mechanisms underlying the dynamic and combinatory distribution across age groups for optimal vaccination outcomes.

## Results

### Model

#### Model construction

In our model, people are classified into four subgroups according to their states: S (susceptible), V (vaccinated), I (infected), and R (removed) (Fig. 1a). The S state means that the corresponding individuals have not yet been infected (including those who have been vaccinated but have not yet gained immunity). The individuals in the V state are those who have been vaccinated and are protected with immunity (we assume that they are not involved in the disease transmission process). We consider the asymptomatic, symptomatic and quarantine phases of COVID-19 as one single I state. The R state consists of recovery and death, and is defined as when an infected individual cannot carry and spread the virus. To accurately describe the virus spreading dynamics, we consider three transition processes between different states: (i) vaccination process (from S to V), in which susceptible people acquire immunity to the disease certain days after vaccination (purple dashed arrow in Fig. 1a); (ii) the infection process (from S to I), in which the state the susceptible individual converts into I state (red dashed arrow in Fig. 1a) while being infected by another infector (red solid arrow in Fig. 1a); and (iii) removal process (from I to R), in which the infected individuals are removed from the virus spreading dynamics due to recovery or death (grey dashed arrow in Fig. 1a). In our model, the infection and removal processes are considered in a non-Markovian manner that the infection and removal rates are time-variant (Fig. 1b).

**Fig. 1.**
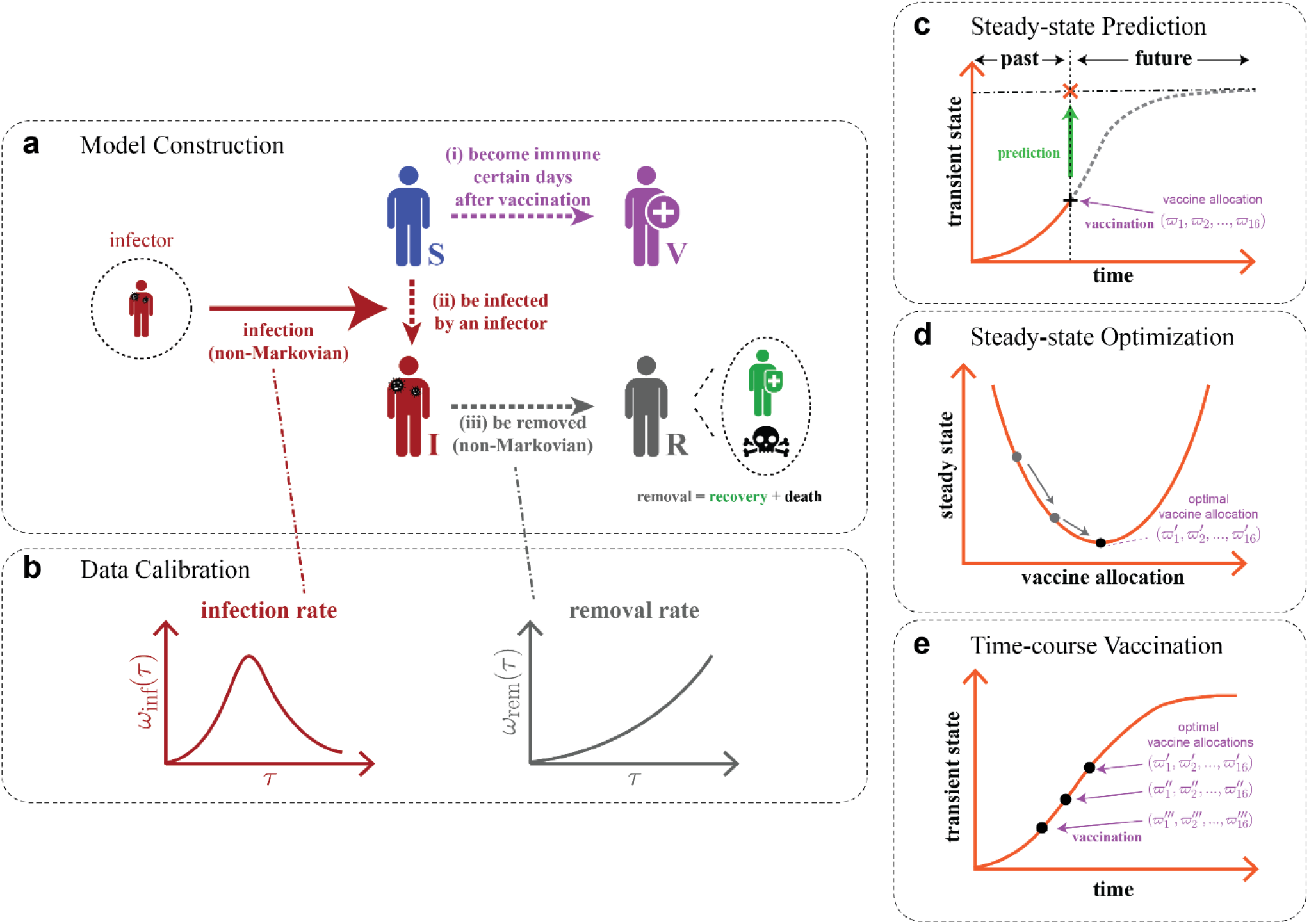
Schematics of optimal vaccine prioritization based on non-Markovian modelling framework. (**a**) Model construction: the states of one individual are composed of S (susceptible), V (vaccinated), I (infected), and R (removed) states (recovery and death); S state can covert to V state several days after vaccination or switch to I state while being infected by another infector in a non-Markovian manner with a time-variant infector infection rate illustrated in (b). I state can turn into R state in a non-Markovian manner with a time-variant removal rate illustrated in (b) due to recovery or death. (**b**) Data calibration: the infection and removal rates and other model parameters (not presented in this figure) are calibrated based on clinical and epidemiological data. (**c**) Steady-state prediction: given a one-off vaccine allocation (denoted by a vaccine distribution vector 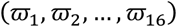 to the 16 age groups at current time point (marked as black “+”), the steady state (marked as orange “×”) can be predicted based on the past epidemic development. (**d**) Steady-state optimization: based on the steady-state prediction in (c), the optimal one-off vaccine allocation that minimizes the desired metric (cumulative infections, cumulative deaths, or years of life lost) can be identified. (**e**) Time-course vaccination: at each vaccination time point, the optimal allocation based on steady-state optimization in (d) is implemented.

The whole population of one country is divided into 16 age groups, including 0–4, 5–9, …, 70–74, and 75+, whose proportions are denoted a vector *p* (Supplementary Information, Section 1.1) [24]. The contact pattern within and between age groups is described by a contact matrix: since the contact pattern during the pandemic is largely affected by the government intervention policy and the public response, we introduce a modulation factor *k* to adjust the unit contact matrix *H*, which means that the contact matrix can be characterized by *kH* (Supplementary Information, Section 1.2 and 1.4) [25].

The spreading dynamics in our model can be described by the following equation:

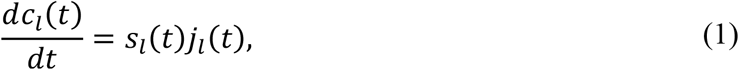

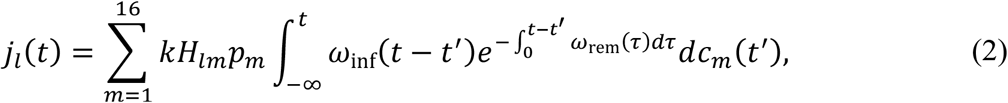

where *c*_*l*_(*t*) and *s*_*l*_(*t*) respectively denote the cumulative infection proportion and susceptible proportion in the age group *l* at time *t*, and *j*_*l*_(*t*) is the infection rate of susceptible population in age group *l* at time *t*. The infection rate *ω*_inf_(*τ*) that an infector transmits disease to a susceptible individual depends on the time duration *τ* starting from the infection of the infector (Supplementary Information, Section 1.3). The removal rate *ω*_rem_(*τ*) that an infected individual is removed due to recovery or death depends on the time length *τ* since the infection of the individual, and 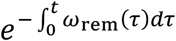 represents the probability that an infected individual has not been removed *t* days after infection (Supplementary Information, Section 1.3). Therefore, these two equations characterize the infection of susceptible population through contacts with infected but not yet removed population in different age groups. It should be noted that, although we here assume a linear adjustment of the unit contact matrix, *H*, by an overall modulation factor *k*, other adjustments of particular age populations, such as school closures, can also be considered in our modelling framework.

The vaccination process affects the time evolution of the susceptible proportion *s*_*l*_(*t*), which can be expressed as follows:

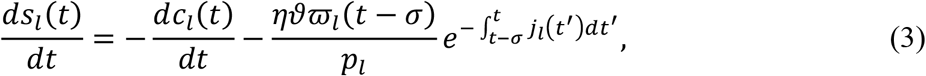

where *σ* denotes the delay between the vaccination and immunity, *η* is the vaccine efficacy, and 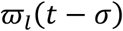 is the vaccine allocation in age group *l* at time *t* − *σ, ϑ* represents the vaccine rollout speed (Supplementary Information, Sections 1.5 and 1.6) [2, 26, 27]. On the right-hand side of Eq. (3), the first term presents the decrease of susceptible population due to infection, and the second term describes that the susceptible people who are vaccinated at time *t* − *σ* are not infected in the following *σ* days, and acquire immunity at time *t*. Note that 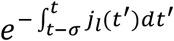 is the probability that a susceptible individual is not infected from *t* − *σ* to *t*.

In our analysis, reducing death and YLL is also important for the vaccine allocation optimization and their evolutions over time are calculated as follows:

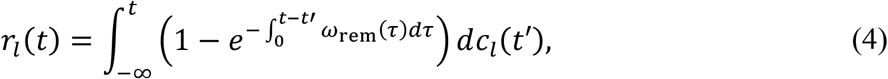

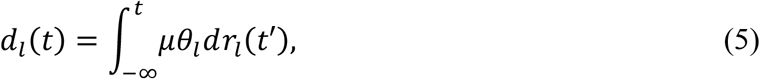

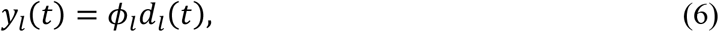

where *r*_*l*_(*t*), *d*_*l*_(*t*), and *y*_*l*_(*t*) are the cumulative removal proportion, cumulative death proportion, and cumulative YLL in age group *l* at time *t*, respectively. The death proportion is strongly related to the infection fatality rate (IFR) distribution: since IFR can be affected by the medical condition and hospital capacity during the pandemic, we introduce a constant *μ* to adjust the unit IFR, *θ*, for the calculation of the cumulative death proportion, which means that the IFR of age group *l* can be calculated as *μθ*_*l*_ (Supplementary Information, Section 1.7) [28, 29]. The average YLL of a death in age group *l, ϕ*_*l*_, is equal to the life expectancy minus the mean age of the age group (Supplementary Information, Sections 1.8) [30].

#### Data calibration

To describe the spreading process more accurately and to make the vaccination strategy more realistic, a series of parameters in our model are all based on real data (Table 1) [24–30]. Some parameters that cannot be collected directly (Fig. 1b), such as infection rate *ω*_inf_(*τ*), removal rate *ω*_rem_(*τ*), contact matrix adjustment parameter *k*, and IFR adjustment parameter *μ*, are calibrated against clinical and epidemiological data (Supplementary Information, Section 1). Infection rate *ω*_*inf*_(*τ*) can be generated by the infection time distribution *ψ*_*inf*_(*τ*), which is the calibration result of the convolution of infectiousness and symptom onset distribution (Supplementary Information, Section 1.3) [14–16]. The removal rate *ω*_*rem*_(*τ*) can be calculated by the removal time distribution *ψ*_*rem*_(*τ*). Following common practice in the epidemiological literature, we take *ψ*_*rem*_(*τ*) to be a Weibull distribution with shape parameter *α* and scale parameter 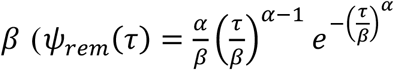, then 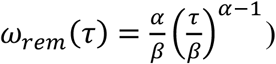, which are estimated by fitting to real data (Supplementary Information, Section 1.3) [31–35]. The calibration of *k* and *μ* after the immunity onset, i.e., *σ* days after the vaccination onset, is not feasible due to the lack of detailed vaccination data of each age group at different time. Therefore, we estimate their values by using the infection and death numbers during a certain time period before the immunity onset (Table 1 and Supplementary Information, Section 1.4). It should be noted that the weakening of epidemic prevention measures during the vaccination period, such as the reopening of school and lifting of travel restriction, can affect the value of *k*, which can also be characterized with our model if related data is available.

**Table 1.**
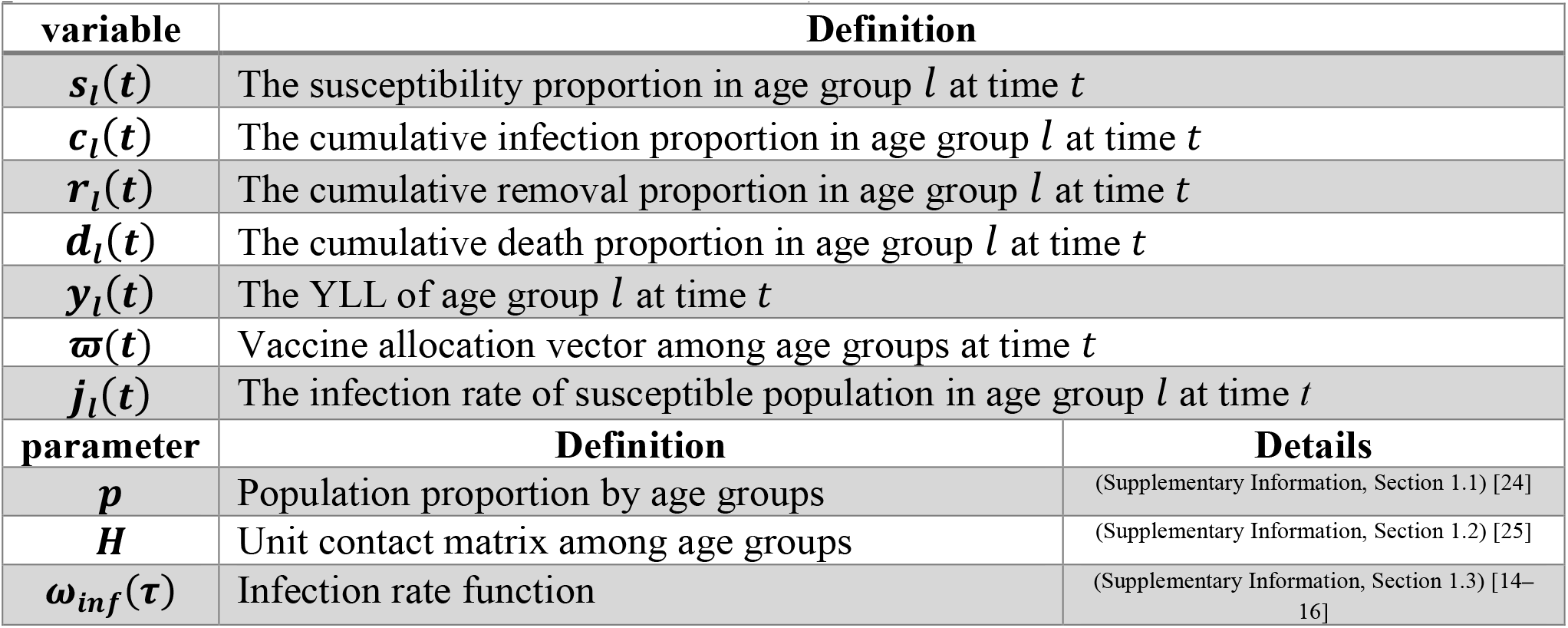
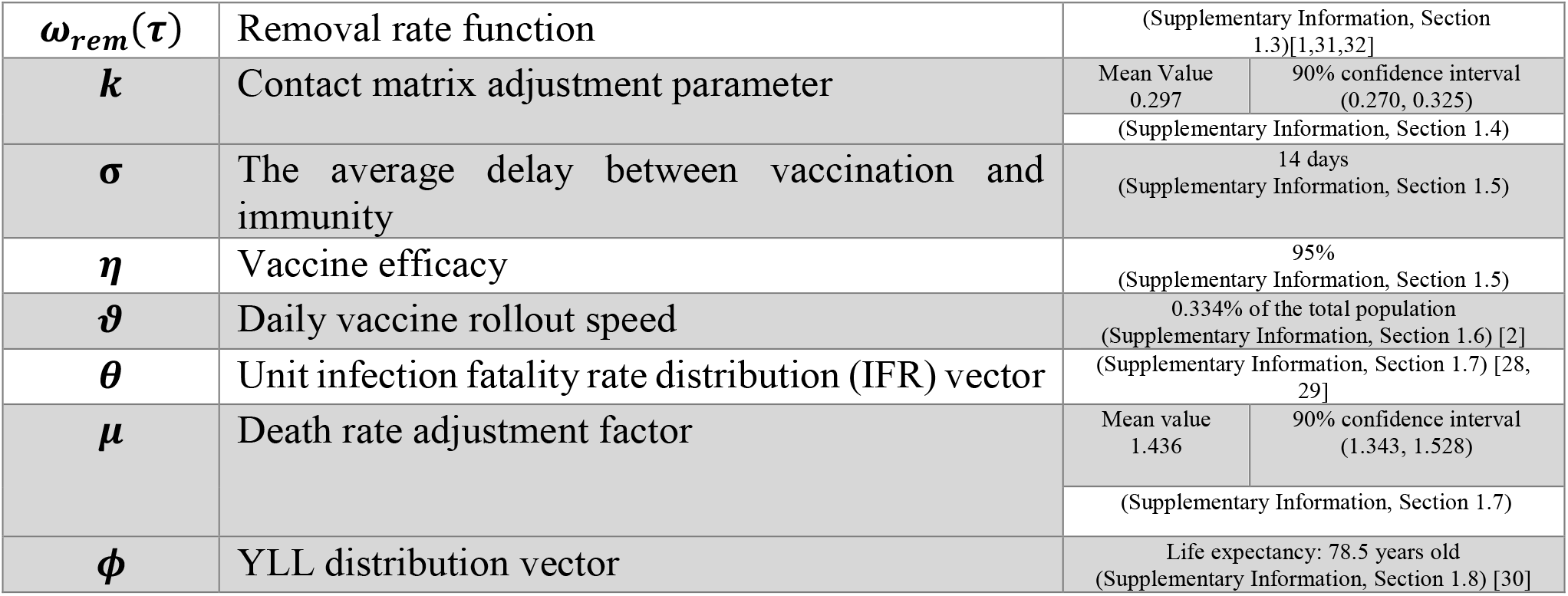
The list of the important variables and parameters (all the calibrated parameter presented in this table are for the United States).

#### Steady-state prediction

Three metrics, including CI (measured as the proportion of cumulative infections in the population), CD (measured as the proportion of cumulative deaths in the population), and YLL, quantify the severity of the pandemic and are the main targets for pandemic prevention. The introduction of vaccines will reduce the susceptible population, which can reduce these three quantities and help to contain the pandemic. Compared to the CI, CD and YLL, final cumulative-infection (FCI), final cumulative-death (FCD), and final years of life lost (FYLL), which are the values of CI, CD, and YLL at the steady state, are more suitable quantities to characterize the efficiency of vaccination strategies, and these three variables are calculated by Eqs. S17–S19 (Supplementary Information, Section 2). In our theoretical framework, if a vaccine distribution is given at one moment, the FCI, FCD, and FYLL can be accurately predicted according to the previous and current non-Markovian conditions, other than by numerically simulating or calculating the total time evolution process approaching the steady state that is computationally expensive (each step of the non-Markovian calculation requires all the current and historical information). Thus, at any moment, there exists a functional relationship between the vaccine allocation and the steady state (Fig. 1c and Supplementary Information, Section 2).

#### Steady-state optimization

To reduce the final loss caused by the pandemic, we utilize the iterative method, sequential quadratic programming, to optimize the steady state based on steady-state prediction [33, 36]. At certain time point, the steady state can be calculated given the vaccine allocation, and optimization enables us to search for the vaccine distribution that can minimize the targeted metric at the steady state (i.e., FCI, FCD, or FYLL) under the constraint of limited daily vaccine supply as illustrated in Fig. 1d (Supplementary Information, Section 2).

#### Time-course vaccination

According to our optimal strategy, the vaccine allocation will be generated by the steady-state optimization at every vaccination time point to form a time-course vaccination strategy. As shown in Fig. 1e, each single vaccine allocation will be optimized to minimize the target metric at the steady state based on the past epidemic development. Note that the optimization at one time point does not consider the following vaccination from that time point, therefore, every single step optimization in our analysis can gain a local optimal effect on the final optimization results.

### Calculations of the pandemic in the United States

In this work, we compared the optimal vaccination strategy (OVS) with several empirical schemes: the pro-rata vaccination strategy (PRVS) which provides a random vaccine distribution, the contact vaccination strategy (CVS) which vaccinates the age group with the highest contact level firstly, the oldest-first vaccination strategy (OFVS) which vaccinates the oldest people firstly, and the youngest-first vaccination strategy (YFVS) which provides vaccines to youngest citizens firstly (Methods: empirical vaccination methods).

We first applied the model to the pandemic in the United States (not consider the effect of mutant strains). In the early stage, the vaccination strategy adopted in the United States was mainly based on people’s occupation and age, and gave priority to older people and those with high-risk occupations [13]. We used real data, including cumulative infection, recovery, and death numbers, in the period from 11 April 2020 to 19 December 2020 (before the vaccination start date 20 December 2020) to calibrate our model by calculating the value of *k* (Supplementary Information, Section 1.4) [1]. According to the model calculation, without vaccination but maintaining similar strictness of pandemic control in the period before vaccination (11 April 2020 to 19 December 2020), approximately 11.392% (i.e., 37.71 million) people will be infected with the novel coronavirus, approximately 0.205% (i.e., 0.68 million) individuals will die, and the FYLL will be 0.040 years (Fig. 2a–c). The United States began its vaccination strategy on 20 December 2020, and on average approximately 0.334% of the population was vaccinated per day between 20 December 2020 and 24 April 2021 (Supplementary Information, Section 1.6) [2]. With this actual vaccination strategy, until 24 April 2021 and 29 June 2021, up to 32.05 and 33.65 million confirmed cases (i.e., 9.682% and 10.167% of the total population), with 0.57 and 0.60 million deaths (i.e., 0.173% and 0.183% of the total population), respectively, were claimed in the United States (Fig. 2a–c) [1,13]. To compare the epidemic prevention effects of different vaccination strategies (OVS, CVS, PRVS, YFVS, and OFVS), we started the vaccination process in our model with the same rollout speed (0.334% of the population per day), the same onset date (20 December 2020), and the end date as 19 December 2021. All vaccine distribution schemes can decrease the FCI, FCD, and FYLL (Fig. 2a–c), and the OVS always performs the best and quickest compared to other strategies (including the real strategies of the United States [13] in terms of minimizing the FCI, FCD, and FYLL, although it generates different dynamic vaccine distributions for different optimization metrics. The OVS can lead the pandemic to reach a steady state rapidly, and the results of 24 April 2021, 29 June 2021, and steady state are extremely close and can be expressed with the same outcomes: With the OVS for reducing FCI, the FCI would reach approximately 7.994% (i.e., 26.46 million people); with the OVS for reducing the FCD, approximately 0.156% (i.e., 0.52 million) individuals would die of the disease; and with the OVS for reducing FYLL, the YLL could decrease to 0.030 years (Fig. 2a–c). Note that OVS generates different vaccine allocations for different optimization metrics, and the vaccine distribution for minimizing FCI may not be the most effective allocation for the other two metrics.

**Fig. 2.**
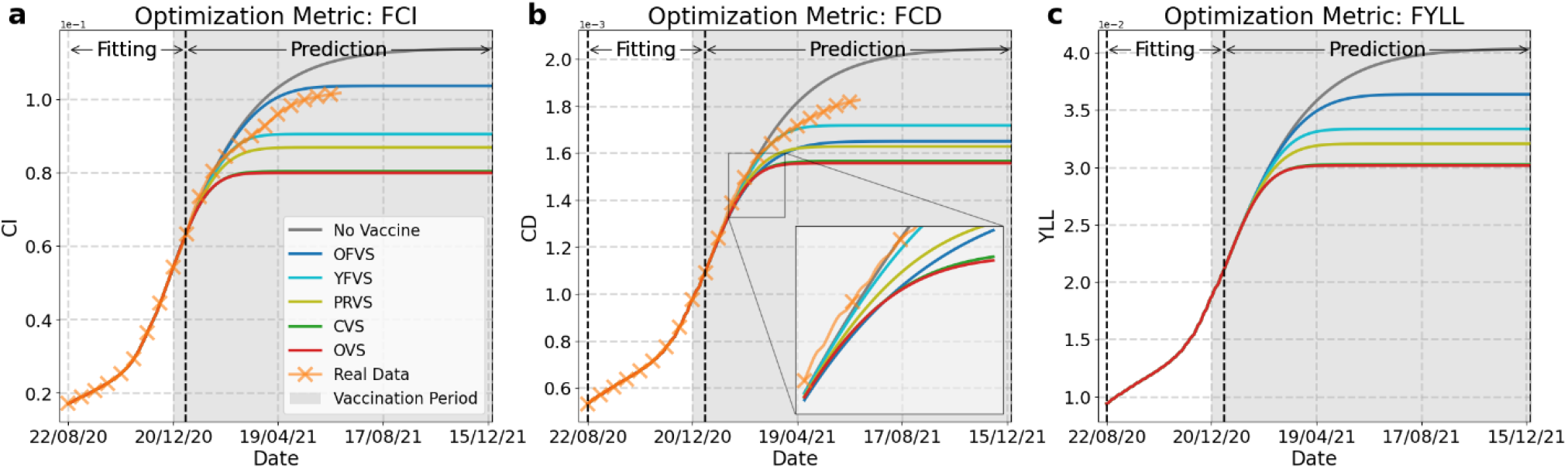
Predictions of COVID-19 spread in the United States under different vaccination strategies. Panels (**a**– **c**), Growth curves of cumulative-infection (CI), cumulative-death (CD), and years of life lost (YLL). The model is calibrated in the ‘fitting’ period (the total ‘fitting’ period starting on 11 April 2020 is too long so is not shown completely) of (a) before immunity onset to estimate the contact matrix adjustment parameter *k*. The real data and calculation values in the ‘fitting’ period of (b) are used to fit the cumulative deaths. The grey areas indicate the implementation of vaccination, and the vertical dashed line in the grey areas denote the onset of immunity 14 days after the first dose of vaccine. We only present the real data until 29 June 2021 because of the impact of the mutant strain (delta variant) in subsequent months.

For the other vaccine empirical strategies, vaccinating the younger first (YFVS) performs better than vaccinating older people firstly (OFVS) in terms of reducing the FCI and FYLL (Fig. 2a and c), while the OFVS is more effectively in reducing the FCD (Fig. 2b) because the IFR is higher in the aged population (Supplementary Fig. 6). Moreover, the PRVS perform better than the OFVS and YFVS in reducing all of the metrics, and the OFVS has a similar reduction effect as the PRVS on the FCD. Vaccinating the individuals with higher contacts first (CVS) performs most closely to our OVS in term of reducing the three metrics (Fig. 2a–c). Although the prevention effect of the CVS is close to that of the OVS in the United States, this does not mean that this phenomenon can also occur in another country such as Germany (Supplementary Information, Section 4). Furthermore, the OFVS has a better prevention effect in reducing the number of deaths than the OVS in a short period (Fig. 2b, inset); however, due to the lack of vaccination for young people, the pandemic will ultimately persist for longer and result in more deaths.

We also predict COVID-19 spreading under different vaccination strategies in other countries, such as Germany and Brazil (Supplementary Information, Section 4). Our method can effectively predict SARS-CoV-2 spreading in these countries over limited time periods as the value of *k* prediction is calculated from the spreading data before vaccination and the fluctuations of *k* after vaccination make it impossible to perform long-term predictions (Supplementary Information, Section 1.4).

### Effect of vaccine availabilities

The availability of vaccines remains a challenging issue in many countries/regions. Although availability is not an issue in the United States, we will study its effect in the model as a control parameter to provide reference for other countries/regions currently lacking vaccines. Vaccine availability is denoted as the percentage of the population who can be maximally covered; thus, this availability corresponds to the time length that the vaccination can continue at a certain rollout rate (0.334% per day in the United States). Subsequently, the model will run without vaccination to reach a steady state, and the values of FCI, FCD and FYLL will be predicted with our theoretical framework. The FCI, FCD, and FYLL with respect to the vaccine availability are presented in Figs. 3a–c. Consistent with intuition, a greater vaccine availability results in a greater reduction in the FCI, FCD, and FYLL in all vaccination schemes. As the availability of vaccines increases, the curves become flatter, indicating that the prevention effects are saturated. Importantly, the results show that the OVS always achieves the optimal epidemic prevention in terms of minimizing the FCI, FCD, and FYLL with any level of vaccine supply availability. The OFVS and YFVS are poor strategies to prevent the pandemic, and the OFVS is better than the YFVS only in reducing the FCD. It should be noted that the OFVS, YFVS, and CVS curves show kinks (Fig. 3a–c), indicating the switch of vaccination between different age groups. When vaccinating a single age group, the prevention effects are best in the initial period, and gradually become ineffective before reaching saturation (Fig. 3a–c) as the age group will play a less important role in the spreading of the disease when the susceptible people are getting fewer. Therefore, vaccinating one or several age group(s) until the individuals in this/these group(s) are vaccinated is not an optimal vaccination strategy for preventing the spread of the virus. To achieve the best prevention effect of the OVS, it is necessary that an elaborately-calculated part of the vaccine distribution be switched to other age groups at certain time points, as will be detailed below.

**Fig. 3.**
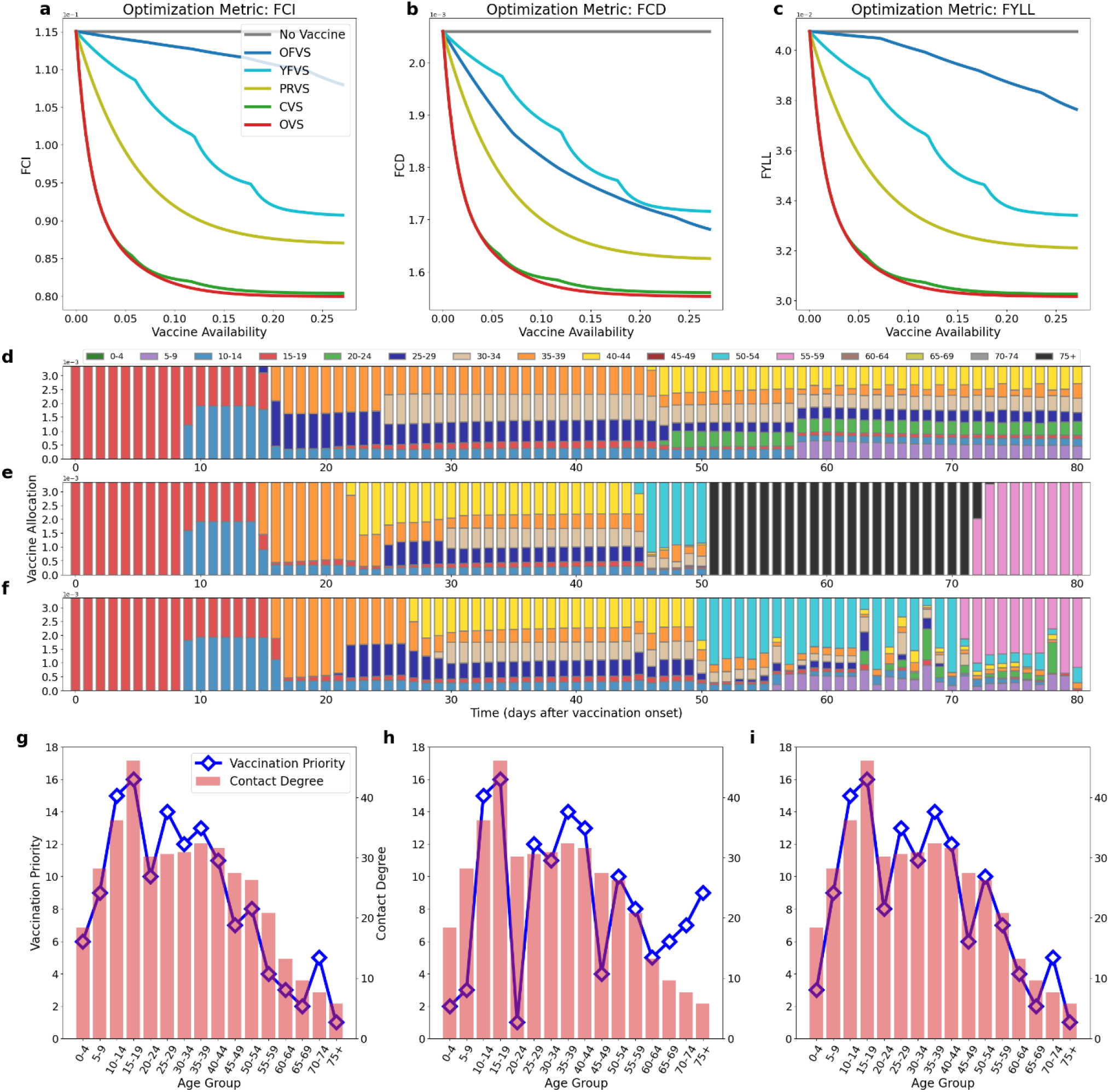
Strategy comparison, vaccine distribution of the optimal vaccination strategy (OVS), and vaccination priorities for age groups with respect to vaccine availability. (**a–c**) The final cumulative-infection (FCI), final cumulative-death (FCD), and final years of life lost (FYLL) under different vaccine availabilities. (**d–f**) Vaccine distribution in different age groups in the OVS with different metrics, FCI, FCD, and FYLL, when the vaccine is available up to given days (horizontal axis). The bar on each single day in the subfigures represents the vaccine distribution to the corresponding age group. (**g–i**) Vaccination priorities for the age groups obtained through the vaccine distributions shown in (d–f) (Methods: calculation of priority), with different targets (FCI, FCD, and FYLL), compared with the contact degree of the age groups in the United States.

Next, we investigated the detailed vaccine distributions in the OVS across different age groups with respect to the vaccine availability (Fig. 3d–f). Given that the vaccine availability can support *n*-day vaccination (with a fixed daily rollout rate), the vaccine distribution of the *n* days is presented as the first *n* bars in Fig. 3d–f. For all three optimization targets, the 15–19-year age group needs to be vaccinated first during the 0–8th day, and the 10–14-year age group should also be vaccinated together with the 15–19-year age group starting from the 9th day. On the 16th day, other age groups need to be included in the vaccination depending on the optimization targets: 35– 39 and 25–29 years for the FCI; 35–39 and 40–44 years on the 22nd day for the FCD; 35–39 and 25–29 years on the 21st day for the FYLL. With the vaccine availability increasing, the 30–34-year age group should follow the 25–29-year age group. The vaccination of the 40–44-year age group should be arranged on the 47th day to minimize the FCI, while it must be advanced to the 22nd and 27th days to minimize the FCD and FYLL, respectively. The biggest difference between the three optimization targets is after the 45–50 days of vaccine availability: to reduce the FCI, the age groups 5–9, 20–24, and 40–44 years need to be vaccinated; to reduce the FCD, the age groups 50–54, 75+, and 55–59 years must be vaccinated in turn; to reduce the FYLL, the age group 50– 54 years should be vaccinated first, followed by simultaneous vaccination of the age groups 5–9 and 55–59 years. The vaccine distributions in the OVS in other countries are shown in Supplementary Information, Section 4.

We next compared the OVS with the CVS to better understand the mechanism of the OVS, as the effects of the OCV and CVS are similar (Fig. 3a–c). The CVS vaccinates different age groups according to their contact degrees, i.e., total contact of an age population (Supplementary Information, Section 1.2), which increase from 0–4 to 15–19 years, suddenly decline from 15–19 to 20–24 years, then increase slowly until 35–39 years, and finally decrease at 40+ years (Fig. 3g– i).

The OVS and CVS have the same prevention effects with limited vaccine availability because both schemes vaccinate the 15–19-year age group first (Fig. 3a–f). For greater vaccine availability, the OVS shows better prevention effects than the CVS. While the CVS only considers the contact degrees of different age groups, the OVS considers all intricate dynamic interactions in the non-Markovian process of virus spreading, which leads to an optimal vaccine distribution.

We obtained the priority of vaccination based on the vaccine distribution in the OVS (Methods: calculation of priority). The vaccination priority to minimize the FCI in the OVS is similar to that in the CVS with small differences (Fig. 3g). Unlike the CVS, which vaccinates different age groups one by one following the contact degree, the OVS vaccinates several age groups simultaneously during certain periods, indicating that only vaccinating an age group with the highest contact in a given period is sub-optimal. The similarity and difference between the contact degree and the optimal priority signifies that the contact level is a main factor in the spreading of the virus and the connectivity pattern between different age groups also has influence on FCI.

For the FCD, the priority of old people increases, while that of people aged 0–4, 5–9, 20–24, and 45–49 years decreases, which can be attributed to the high IFR in older age groups. However, the age groups with the highest priorities are still those with high contact degrees. This result suggests that the contact level is still the main factor influencing the FCD. Therefore, to minimize the FCD, we must consider the integrated influences of transmission and IFR (this will also be further analysed in section ‘Mechanism of dynamic vaccine allocation of OVS’).

For the FYLL, the priority distribution is a compromised version of those for the FCI and FCD. Vaccination of age groups with high contacts remains the top priority. The priority of the age groups 0–4, 5–9, 20–24, and 45–49 years is higher for the FYLL than for the FCD because death in the young population has a greater contribution to the YLL. The priority of old people does not increase with age, even if the IFR is high, as the death of old people contributes less to the YLL.

In addition, we also present how the vaccine efficacy affects the vaccination duration in the United States (Supplementary Information, Section 5). Higher efficacy can result in a faster decrease in new CI, CD, and YLL and a greater reduction in the FCI, FCD, and FYLL by using the vaccination allocations provided by the OVS (Supplementary Figs. S13). The vaccination strategies have different effects in different countries (Supplementary Information, Section 6). In Brazil the vaccination shows the least effects in reducing the FCI and FCD, and in Germany the vaccines have the smallest effects in reducing the FYLL.

### Mechanism of dynamic vaccine allocation of OVS

We have shown the detailed optimal time-course vaccine prioritization for respectively minimizing the FCI, FCD, and FYLL in the United States. However, given the OVS, it remains unclear why the vaccines should be switched dynamically, why combinatory vaccination is needed, and how the virus spreading and vaccination process jointly affect the optimal time-course allocations. Elucidating the vaccination mechanism behind the OVS will enable us to gain insight into the optimal vaccine prioritization strategy and better guide pandemic prevention.

To analyse the time-course mechanism of the OVS, we introduce a new quantity, the prevention efficiency with vaccine (PEV), which is defined as the reduction of target metrics (FCI, FCD, or FYLL) under the assumption that an age group or a combination of age groups is vaccinated with a given number of vaccines (Methods: definition of PEV). At a given time, a distribution scheme with the largest PEV will be the most efficient. In this study, we use the symbols 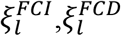, and 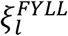 to denote the PEV of the age group or age group combination *l* for reducing FCI, FCD, and FYLL, respectively. Mathematically, the PEV at a given moment is equivalent to the ‘minus difference quotient’ of the predicted steady state with respect to an imaginary vaccination of an age group or a combination of age groups at that moment, while the actual vaccination may not be implemented or can be administered to other populations; thus, the PEV depends on the transient state of the spreading dynamics evolved from previous vaccination history.

In the main text, we focus on the PEV for the target of FCD, and the analyses for FCI and FYLL are presented in Supplementary Information, Section 7. We first implement no vaccination in the virus spreading dynamics course and calculate 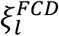 for each of the 16 age groups on each day with the same rollout speed as that in the United States (Supplementary Information, Section 1.6), from the actual vaccination onset, 20 December 2020 (Fig. 4a). With the transmission of disease to a larger proportion of the population, the PEVs of all age groups decline without intersections between different curves, suggesting that vaccination becomes less efficient with further spreading of the disease, while the relative prioritizations of different age groups remain fixed.

**Fig. 4.**
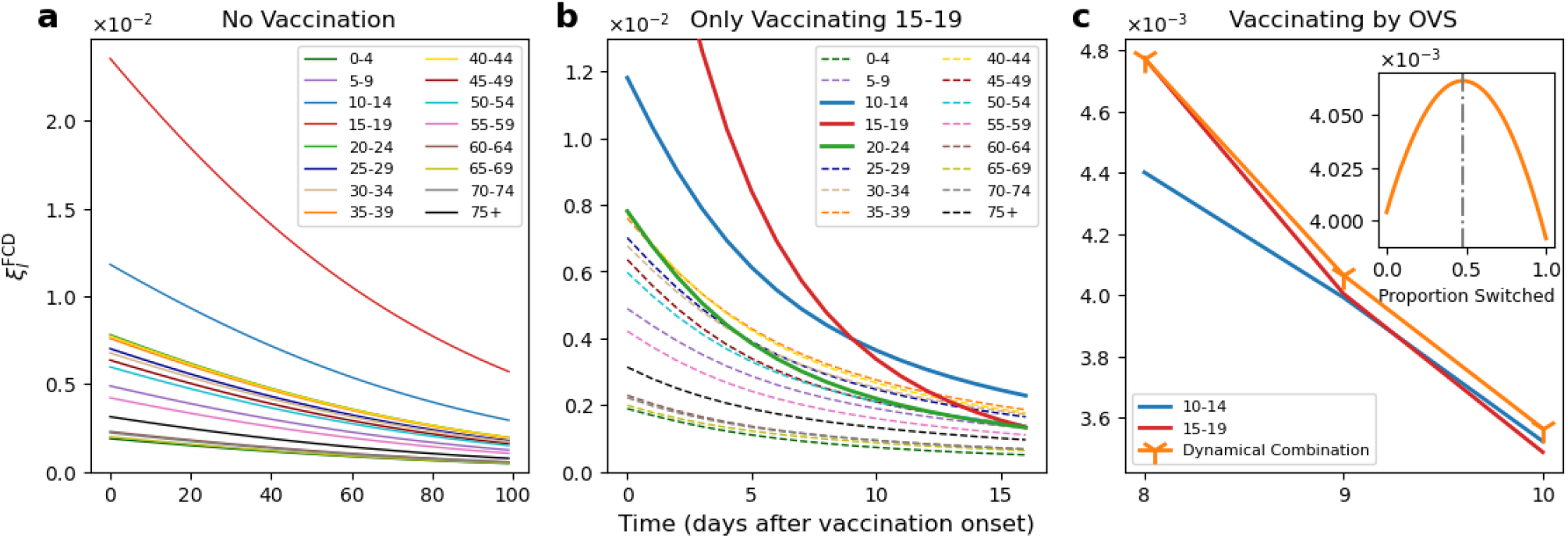
Mechanisms of the optimal vaccination strategy (OVS) for reducing the final cumulative deaths (FCD). (**a**) Synchronous change of prevention efficiency with vaccine (PEV) for reducing the FCD of each age group with the evolution of spreading dynamics when vaccination is not implemented. (**b**) Dynamic changes in the PEV of each age group while the 15–19-year age group is vaccinated. Note that the PEV of 15–19-year age group declines fastest. (**c**) Detailed dynamic changes in the PEVs of the 10–14- and 15–19-year age groups, and their combination. Inset: orange solid line shows the changes in the PEV for reducing the FCD under different vaccine percentages switched from the 15–19-year age group to the 10–14-year age group on the 9th day (0 indicates that all vaccines are provided for the 15–19-year age group, and 1 indicates that all vaccines are supplied to the 10–14-year age group). The vertical grey-dashed line passing the maximum represents the percentage generated by the OVS as shown in Fig. 3e.

If we vaccinate a single age group (15–19 years) each day with the same rollout speed as that in the United States from 20 December 2020, the PEVs of the vaccinated group and all the other age groups will be affected, and those groups with higher contacts with the vaccinated group will be more strongly influenced (Fig. 4b, red, blue, and green lines). The difference among the dynamic PEV changes of different age groups caused by the inhomogeneous contact results in intersections of the PEV curves. For example, the PEV of the 15–19-year age group becomes smaller than that of the 10–14-year age group when continuing vaccination of the 15–19-year age group to the 9th day (Fig. 4b). This suggests that further vaccinating this 15–19-year age group is no longer the most efficient strategy. Therefore, a vaccination switch around the curve intersection is necessary to achieve optimal vaccine allocation.

Interestingly, the vaccination switch does not simply involve switching all of the vaccines to the age group with the highest PEV. When we implement the OVS, the vaccine allocations during the 0–8th days are only to the 15–19-year age group (Fig. 3d–f) because the PEV of the 15–19-year age group during this period is much higher than those of other age groups (Fig. 4b). However, when the PEVs of the 15–19- and 10–14-year age groups become close, only vaccinating the 15– 19-year age group is no longer the optimal allocation (Fig. 4c). On the 9th day of vaccination, vaccinating the 10–14- and 15–19-year age groups simultaneously is the best prevention strategy (Fig. 4c, orange line). Moreover, the vaccine allocation ratio between the two populations needs to be adjusted according to the OVS to optimize the prevention effect (Fig. 4c inset). Therefore, we reveal that the dynamic variation of the PEV and the change of ranking among the age groups caused by continuous vaccination in an inhomogeneous contact network is the main reason for the OVS to switch vaccines from one age group to another with a dynamic combination of vaccines. It should be noted that the PEV curve of the 20–24-year age group intersects many lines of other age groups to shift to lower priority while vaccinating the 15–19-year age group; this explains why the 20–24-year age group has extremely low priority for vaccination (Fig. 3h). The same analysis and conclusions apply to the FCI and FYLL (Supplementary Information, Section 7 and Supplementary Figs. S15–S16).

## Discussion

We propose an optimal age-stratified vaccination strategy to minimize important metrics (FCI, FCD, or FYLL) based on steady-state prediction in the data-calibrated non-Markovian spreading dynamics. According to our results, we can minimize the losses caused by the COVID-19 pandemic and maximize the effect of vaccines on epidemic prevention by allocating vaccines dynamically to different age groups. The results obtained using our model differ from those of several previously published works, including Bubar *et al*. [3], Matrajt *et al*. [4], and Buckner *et al*. [5]. Bubar *et al*. compared a few pre-determined schemes, each covering broad age ranges, and showed that vaccines should be prioritized to adults aged 20–49 years to reduce the cumulative incidence, and that allocating vaccines to individuals older than 60 years can minimize the deaths and YLL [3]. Matrajt *et al*. illustrated that to minimize the deaths and YLL, an OVS in the Markovian framework will need to either distribute vaccines to those with high IFRs or allocate vaccines to those with high transmission rates [4]. Buckner *et al*. considered the effect of essential workers and found that to impede spreading, the optimal policies should prioritize vaccination to essential worker groups with a high risk of infection, while first vaccinating those with a high risk of fatality is the optimal strategy in controlling the number of deaths [5]. In contrast, according to our analysis, although vaccination priority of senior people with high fatality needs to be enhanced to minimize deaths, they should not be prioritized over people with a high risk of transmission; therefore, vaccines should be prioritized to the groups with a high risk of infection due to a high contact degree for controlling all the three metrics (FCI, FCD, and FYLL). Prioritizing older people ignores impeding the transmission of viruses and ultimately results in more deaths than the OVS, although this strategy can reduce cumulative deaths in a short period [5].

These differences mainly result from the different model constructions, parameter selections, and optimization methods. The previous three studies only considered the memoryless Markovian spreading dynamics, while our analysis uniquely improves the model to a non-Markovian framework that fully considers the temporal sequences of the occurrences of the key events, including infection, removal, and vaccination. While the selected parameters in our work and the previous three studies are all based on certain data values, in reality, the temporal characteristics of the dynamics require a detailed description from the probability density function of the stochastic process. For example, in our model, the non-Markovian infection mostly occurs around the symptom onset according to clinical data, and can overcome the limitations of Markovian assumption characterized by constant infection rates during different symptom periods, which can underestimate the impact of transmission. In addition, optimization approaches and metrics are essential to identify the groups to be prioritized for vaccination. Matrajt *et al*. and Bubar *et al*. considered static optimization, wherein the vaccine allocation and administration strategies did not change over time [3, 4]. Similar to Buckner *et al*. [5], our optimization is also dynamic, adjusting for changing epidemiological transmission over a period; however, our optimization target of each vaccine allocation adjustment is to minimize the corresponding metrics in steady state, while Buckner *et al*. focused on optimizing the objectives in the transient state. Because vaccinating old people can reduce the cumulative deaths effectively in the initial stage of vaccination, as we demonstrated, optimizing a short-term prevention effect in reducing the deaths may entail vaccinating older essential workers, which was the conclusion made by Buckner *et al*.

While the optimal vaccine distribution strategies differ across countries as the model is calibrated by actual situations, fortunately, we can also elucidate some common features among the vaccination strategies of the three countries, i.e., the United States, Germany and Brazil. To minimize the three metrics, vaccinating people with higher contacts first to block the virus transmission is the most important measure; meanwhile, to minimize the FCD, older people should be allocated vaccines after the spread of disease has been contained to a certain extent.

Our theoretical analysis of the mechanisms of the time-course vaccine allocation optimization reveals that continuous vaccination will lead to a decline in the efficiency of vaccination, suggesting that earlier vaccination is both more effective and more desirable. More importantly, in inhomogeneous networked populations, continuous vaccination can induce intricate dynamic variation in the PEV and the relative ranking across age groups, which in turn induce dynamic changes in the vaccination prioritization of age groups. This type of mechanism prompts the dynamic vaccine switch and combinatory vaccine allocation for the optimal strategy. While previous work [5] has considered the time-course optimization, how previous optimal vaccine allocations affect future optimization and what leads to the key characteristics of a dynamic and combinatory optimal time-course vaccination strategy have not been studied and analysed. Our current work has successfully addressed this issue, which facilitates a deeper understanding of time-course vaccination optimization.

Our analysis provides a significant reference for future prevention policy making in countries that still lack sufficient vaccine supply. Moreover, the modelling framework can be extended to the vaccination strategy to control future spreading of mutant strains. However, some matters still require attention. As our optimal vaccine allocation can change every day, this may lack feasibility for actual implementation. However, this extremely detailed vaccine distribution can be adjusted by summing the vaccine distributions during a certain period. For example, if the government must implement an unchangeable vaccination strategy during the first month, they can add all of the model-predicted vaccine distributions of the first 30 days and obtain the total vaccine allocation proportions for all age groups as the allocation strategy to be utilised for the 1-month vaccination. In reality, there will be other limiting factors for the prioritization of vaccines, e.g., the different risks of side-effects in different populations, and such constraints could be incorporated into our optimization schemes.

## Methods

### Empirical vaccination methods

PRVS always gives a vaccine distribution proportional to the susceptible population in every age group at that moment. CVS and OFVS will list all age groups in descending order by contact rate and age, respectively. In contrast, YFVS lists them in ascending order by age. Then every group will be vaccinated until the people in the group ranked above are vaccinated; for example, the second group can only be vaccinated after all of the people in the first group are vaccinated. For all vaccination strategies, the sum of the vaccines distributed to all the age groups equals the vaccine supply at that moment.

### Calculation of priority

Priorities vary with the change in the evaluation standard. For simplicity, we assume that the age groups that get vaccinated earlier have the higher priorities. We ranked the 16 age groups in descending order according to the order of their vaccination (where 16 is the population that was the first to be vaccinated), and their rank order in the list is taken as their priority.

### Definition of PEV

To calculate the PEV of one age group or an age group combination *l* at a certain time point *t*_0_ and with a certain vaccine amount *ϑ*, we must obtain the prediction of FCI, FCD, or FYLL, denoted as 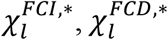 or 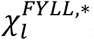, respectively, if we add the vaccine to the age group or age group combination *l*. Meanwhile, we can also calculate the prediction of FCI, FCD, or FYLL, denoted as 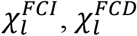, or 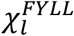, respectively, if we add no vaccine to any age group. Then, the PEV of the age group or age group combination *l* for reducing the FCI, FCD, and FYLL, denoted as 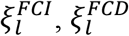, and 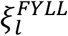, respectively, can be calculated from the reduction rates as follows:

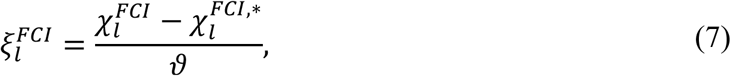

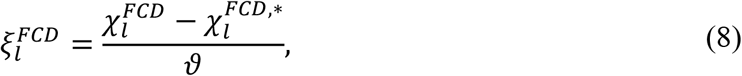

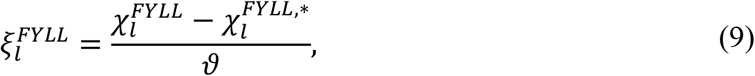

which can reflect the effect of vaccinating the age group or age group combination with the given amount of a certain vaccine. And Eqs. (7–9) can be regarded as minus difference quotients of the functions of steady state with respect to vaccine allocation.

## Supporting information

Supplemental Tables, Figures and Description

## Data Availability

All data produced in the present study are available upon reasonable request to the authors

## Acknowledgements

This work was supported by the Hong Kong Baptist University (HKBU) Strategic Development Fund. This research was conducted using the resources of the High-Performance Computing Cluster Centre at HKBU, which receives funding from the Hong Kong Research Grant Council and the HKBU. Competing interests: The authors declare that they have no competing interests. Data and materials availability: All data needed to evaluate the conclusions in the paper are present in the paper and/or the Supplementary Information. Additional data related to this paper may be requested from the authors.

## Contributions

M.F., L.T. and C.-S.Z. designed research; M.F. performed research; L.T. and C.-S.Z. contributed analytic tools; M.F., L.T. and C.-S.Z. analysed data; M.F., L.T. and C.-S.Z. wrote the paper.

## Notes

### Competing Interest Statement

The authors have declared no competing interest.

### Author Declarations

https://systems.jhu.edu/research/public-health/ncov https://ourworldindata.org/covid-vaccinations https://population.un.org/wpp https://www.mscbs.gob.es/profesionales/saludPublica/ccayes/alertasActual/nCov/documentos/Actualizacion_103_COVID-19.pdf https://apps.who.int/gho/data/node.main.688

