## Supplemental Tables, Figures and Description for "Mechanism of optimal time-course COVID-19 vaccine prioritization based on non-Markovian steady-state prediction"

**2 Steady-state Prediction** **8**

**3 Steady-state Optimization** **10**

**4 Pandemics in Other Countries** **11**
4.1 Pandemics in Germany . . . . . 11
4.2 Pandemics in Brazil . . . . . 12

**5 Vaccination Duration with Different Efficacies** **15**

**6 Optimal Vaccination Effects in Different Countries** **16**

**7 PEV for Reducing FCI and FYLL** **17**

*
†

### Section 1: Model parameters

#### Section 1.1: Populations Grouped by Age

In our analysis, the total population (from 0 to 100 years old) has been divided into 16 different groups according to people's ages. Each of the first 15 age groups consists of five consecutive ages (i.e., 0–4, 5–9,..., and 70–75), while the last age group is composed of the ages from 75 to 100 (i.e., 75+). The figures of age population distributions in the United States, Germany, and Brazil are presented (Supplementary Fig. 1a–d) [1]. Note that the individuals from 75 to 100 years old are classified into the age group 75+, so the bar of this group is much higher than others.

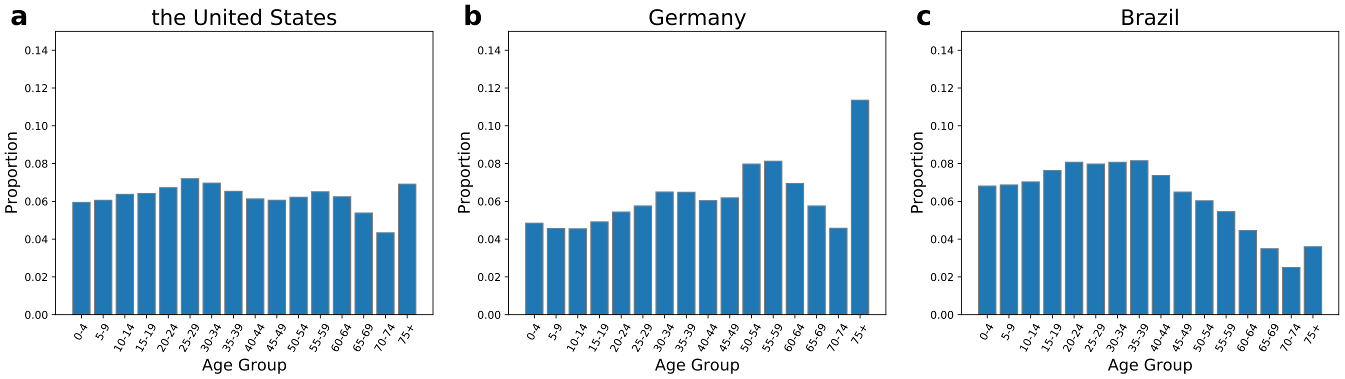

Supplementary Fig. 1: **Age distributions of three countries.** **a** the United States; **b** Germany; **c**. Brazil.

#### Section 1.2: Contact Matrix and Contact Vector

The unit contact matrix, whose $lm$th element denotes the mean number of contacts between age group $l$ and age group $m$, can mathematically characterize the contacts within and between different age groups (Supplementary Fig. 2a–d) [2]. The bright diagonals indicate that people mainly contact people of a similar age. The other two off-diagonal bright zones on both sides of the diagonals reflect the contacts among families, and the home contacts in Germany occupy a big proportion in contrast to other countries.

In addition, the original data of contact matrix $\hat{H}$ in Ref. [2] only describe the directional contacts from contactors to contactees. However, contacting others and being contacted can both facilitate the transmission of viruses. So to deal with this issue, we obtain a symmetrical contact matrix $H$ by adding $\hat{H}$ and its transposed matrix $\hat{H}^T$,

$$H = \hat{H} + \hat{H}^T \tag{S1}$$

Moreover, in order to characterize one age group's ability of contacting with other groups, we define the contact vector of age group $l$ as

$$h_l = \sum_{m=1}^{16} H_{lm} \tag{S2}$$

where $l$ and $m$ are the indexes of the 16 age groups, and every element in the contact vector is defined as contact degree while each element of the contact matrix is called contact rate. The average contact degree of Brazil is the highest in the three countries while that of Germany remains the lowest (Supplementary Fig. 2e–h). For the people aged 0 to 19 in the three countries, contact degrees increase with age. But the contacts drastically decline when individuals grow up to 20–24 years old, likely because the people in this age group have just graduated from universities and lose a great deal of their social relations developed in campuses. And then contacts rise again, peak at 35–39 (for the United States) or 40–44 years old (for Germany) and finally decrease as the age increases. But an exception is that the contact degree of Brazil declines all the way when the age gets greater than 19. Moreover, although different contact patterns exist in different nations, they still present a few characteristics similarities. For instance, teenagers always behave significantly actively, but their social relations are limited to peer contacts. In contrast, middle-aged people have more widespread social connections.

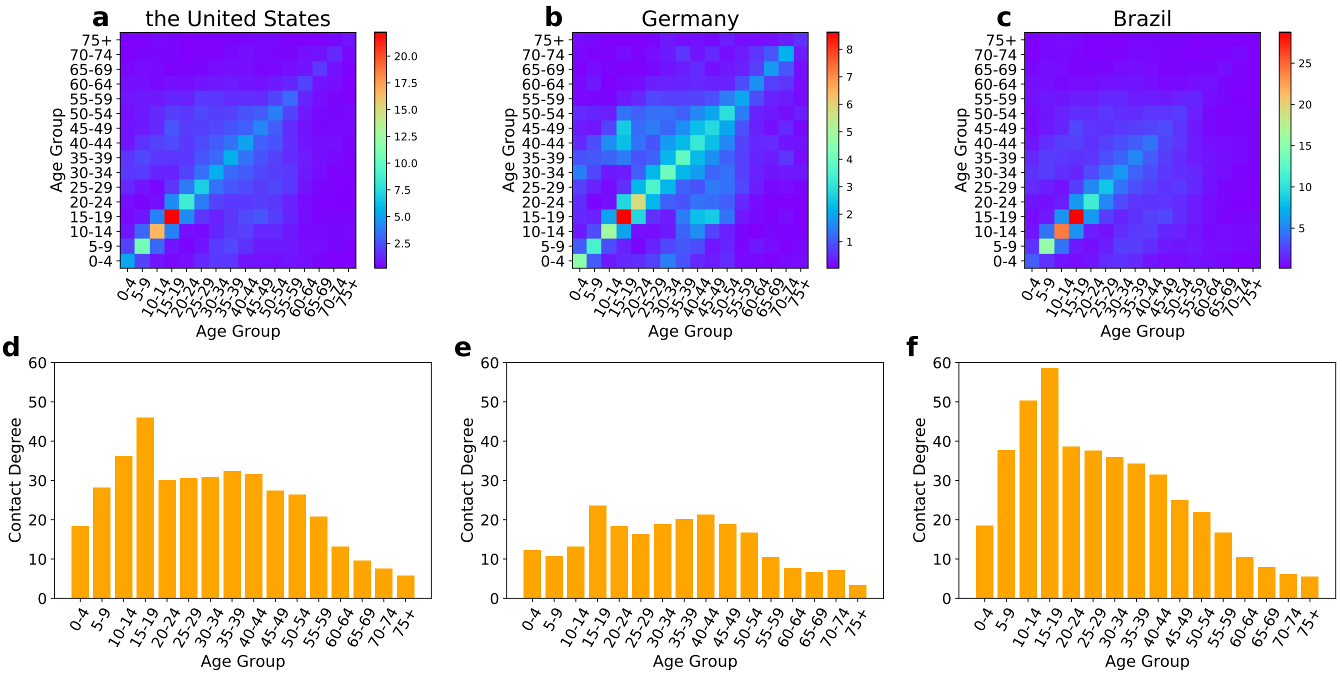

Supplementary Fig. 2: **Unit contact matrices and contact vectors in three countries.** Upper panels **a**–**c** the unit contact matrices of the United States, Germany and Brazil. Lower panels **d**–**f** the contact vectors of the United States, Germany and Brazil.

#### Section 1.3: Infection and Removal Time Distribution

##### 1.3.1 Definitions and Physical Meanings

To fully describe the non-Markovian spreading process, we defined a stochastic variable $T$ of an infected individual as the time duration between the onset of being infected and the current time. For example, if the infected individual was infected at the time $t'$ and the current time is $t$, then $T = t - t'$. We can divide the whole process of this compartmental model into three parts as follows: a susceptible individual is infected by another infected individual and the probability of being infected by this virus host with $T \in [\tau, \tau + d\tau)$ equals to $\omega_{\text{inf}}(\tau)d\tau$, where $\omega_{\text{inf}}(\tau)$ is the time-dependent infection rate; an infected person with $T \in [\tau, \tau + d\tau)$ can be removed from the spreading process with a certain probability $\omega_{\text{rem}}(\tau)d\tau$, where $\omega_{\text{rem}}(\tau)$ is the time-dependent removal rate. Therefore, non-Markovian infection and removal processes can be fully described by the infection and removal rate functions, $\omega_{\text{inf}}(\tau)$ and $\omega_{\text{rem}}(\tau)$. The infection time distribution $\psi_{\text{inf}}(\tau)$ and the removal time distribution $\psi_{\text{rem}}(\tau)$ can also be characterized as the infection and removal processes, respectively, which can be deduced as follows:

$$\psi_{\text{inf}}(\tau) = \omega_{\text{inf}}(\tau)e^{-\int_0^\tau \omega_{\text{inf}}(\tau')d\tau'} \tag{S3}$$

$$\psi_{\text{rem}}(\tau) = \omega_{\text{rem}}(\tau)e^{-\int_0^\tau \omega_{\text{rem}}(\tau')d\tau'} \tag{S4}$$

The physical meanings of these two distributions are illustrated as follows: $\psi_{\text{inf}}(\tau)d\tau$ denotes the infinitesimal probability that the $T$ of an infected individual, when transmitting the virus to a certain susceptible person, is in the interval $[\tau, \tau + d\tau)$; $\psi_{\text{rem}}(\tau)d\tau$ denotes the infinitesimal probability that the $T$ of an infected individual, when they recover from or die of the disease, stays in the interval $[\tau, \tau + d\tau)$.

Moreover, we can use $\Psi_{\text{inf}}(\tau)$ and $\Psi_{\text{rem}}(\tau)$ to respectively denote the probabilities that the infected individual never transmits the virus and never gets removed (i.e., recovers or dies) during the period $T \in [0, \tau)$, which can be expressed by $\psi_{\text{inf}}(\tau)$ and $\psi_{\text{rem}}(\tau)$ as

$$\Psi_{\text{inf}}(\tau) = 1 - \int_0^\tau \psi_{\text{inf}}(\tau')d\tau' \tag{S5}$$

$$\Psi_{\text{rem}}(\tau) = 1 - \int_0^\tau \psi_{\text{rem}}(\tau')d\tau' \tag{S6}$$

Although $\omega_{\text{inf}}(\tau)$, $\psi_{\text{inf}}(\tau)$, and $\Psi_{\text{inf}}(\tau)$ (or $\omega_{\text{rem}}(\tau)$, $\psi_{\text{rem}}(\tau)$, and $\Psi_{\text{rem}}(\tau)$) have different physical meanings, they all characterize the same process and any of them can be deduced by any other.

###### 1.3.2 Calibration of Infection Time Distribution

In our analysis, we assume that all the age groups of all the countries share the same unit infection rate function. According to clinical data, the infectiousness $\psi_{\text{clinic}}(\tau)$ relative to the onset of symptoms can be fitted (Supplementary Fig. 3a) [3–5]. The infection peaks around the symptom onset, and the decline after symptom onset is mainly because the patients with symptoms will undergo compulsory quarantines. Moreover, the symptom onset time after being infected can also be characterized by a time distribution $\psi_{\text{sym}}(\tau)$ and the symptom mostly appears about 4 days after contracting the virus (Supplementary Fig. 3b) [5]. And then, the infection time distribution $\psi_{\text{inf}}(\tau)$ related to the unit infection rate function can be calculated by Eq. S7 (Supplementary Fig. 3c),

$$\psi_{\text{inf}}(\tau) = \int_0^{+\infty} \psi_{\text{clinic}}(\tau - \tau')\psi_{\text{sym}}(\tau')d\tau'. \tag{S7}$$

The infection time distribution can vary over time and $\psi_{\text{inf}}(\tau)$ shown in Supplementary Fig. 3c can only reflect the infection process for a short period. However, under our assumption, all the values of the corresponding infection rate function $\omega_{\text{inf}}(\tau)$ can develop proportionally, meaning that the ratio of the values at every two $\tau$ always keeps constant.

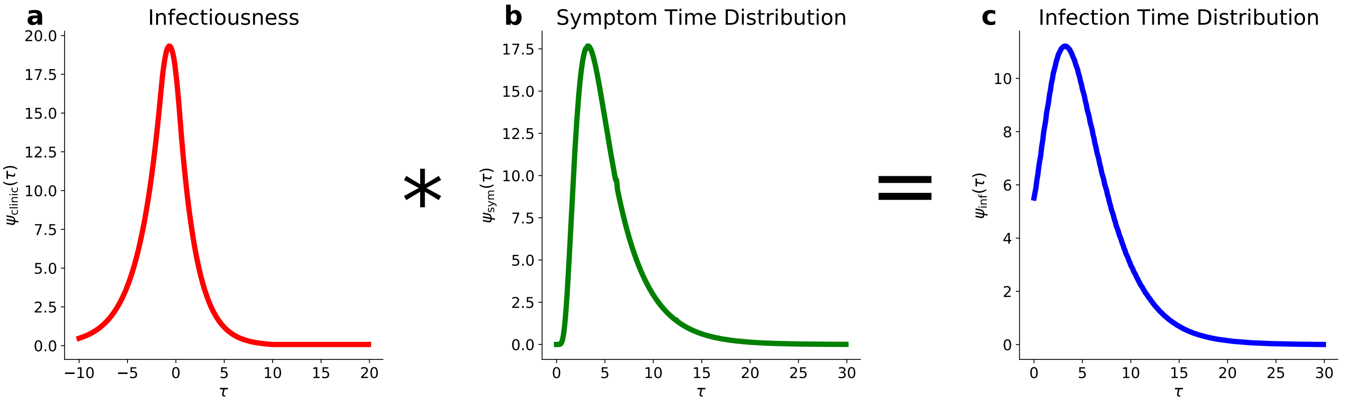

Supplementary Fig. 3: **Infection time distribution calculated by the convolution of infectiousness and symptom onset distribution.** **a** the curve of infectiousness; **b** symptom onset distribution; **c** infection time distribution related to the unit infection rate function.

###### 1.3.3 Calibration of Removal Time Distribution

The removal process is composed of recovery and death processes meaning that the infected individual is removed from the disease transmission and no longer spreads disease if he/she recovers or dies. This combined process is always related to medical conditions and epidemic prevention measurements of one country, which are not easy to be quantified by certain parameters or clinical data. Therefore, a Weibull assumption of removal time distribution and numerical fitting by real cumulative-infection (cumulative-infection is defined as the proportion of cumulative infections in total population) and current-infection (current-infection is defined as the proportion of current infections in total population) data is becoming a preferred approach [6]. According to the following equations,

$$\psi_{\text{rem}}(\tau) = \frac{\alpha_{\text{rem}}}{\beta_{\text{rem}}}\left(\frac{\tau}{\beta_{\text{rem}}}\right)^{\alpha_{\text{rem}}-1} e^{-\left(\frac{\tau}{\beta_{\text{rem}}}\right)^{\alpha_{\text{rem}}}}, \tag{S8}$$

$$i_{\text{calc}}(t) = \int_{-\infty}^{t} \Psi_{\text{rem}}(t-\tau)dc_{\text{real}}(\tau), \tag{S9}$$

where $c_{\text{real}}(t)$ is the real cumulative-infection according to data from Ref. [6], and $i_{\text{calc}}(t)$ is the calculated current-infection. According to Eq. S8–S9, we define a lose function as

$$L(\alpha_{\text{rem}}, \beta_{\text{rem}}) = \frac{\int_0^t [i_{\text{calc}}(t') - i_{\text{real}}(t')]^2 dt'}{t} \tag{S10}$$

where $i_{\text{real}}(t)$ is the real current-infection with respect to $t$ [6]. Therefore, the appropriate $\alpha_{\text{rem}}$ and $\beta_{\text{rem}}$, based on Eq. S9 which can lead $c_{\text{real}}(t)$ to calculate the $i_{\text{calc}}(t)$ closest to $i_{\text{real}}(t)$ (i.e., minimizing the loss function Eq. S10), of different countries can be obtained by trust-region constrained method (Supplementary Table 1) [7–9]. In addition, the corresponding removal time distributions are also presented (Supplementary Fig. 4). In the parameter fitting, the data about $c_{\text{real}}(t)$ and $i_{\text{real}}(t)$ are selected from 16 June 2020 to 15 October 2020 for all the three countries. Because some states/regions of the United States lack the data about recovered cases, we only select the data from the 25 states/regions with complete records (including Wyoming, Wisconsin, West Virginia, Virgin Islands, Vermont, Utah, Texas, Tennessee, South Dakota, Pennsylvania, Oklahoma, Ohio, North Dakota, North Carolina, New Mexico, New Hampshire, Montana, Mississippi, Minnesota, Michigan, Massachusetts, Louisiana, Iowa, Guam, Arkansas) and then add them together as the data of the United States only for the removal time distribution fitting purpose. We also hypothesize that the removal processes of all the age groups in one country are characterized by the same distribution.

| Country | $\alpha_{\text{rem}}$ | $\beta_{\text{rem}}$ |
|---|---|---|
| the United States | 1.93 | 21.67 |
| Germany | 3.44 | 16.22 |
| Brazil | 6.28 | 12.90 |

Supplementary Table 1: **Parameters ($\alpha_{\text{rem}}$ and $\beta_{\text{rem}}$) of Weibull removal time distributions for the United States, Germany and Brazil ($\zeta$ is the average days that infected individual will stay in I state before removal).**

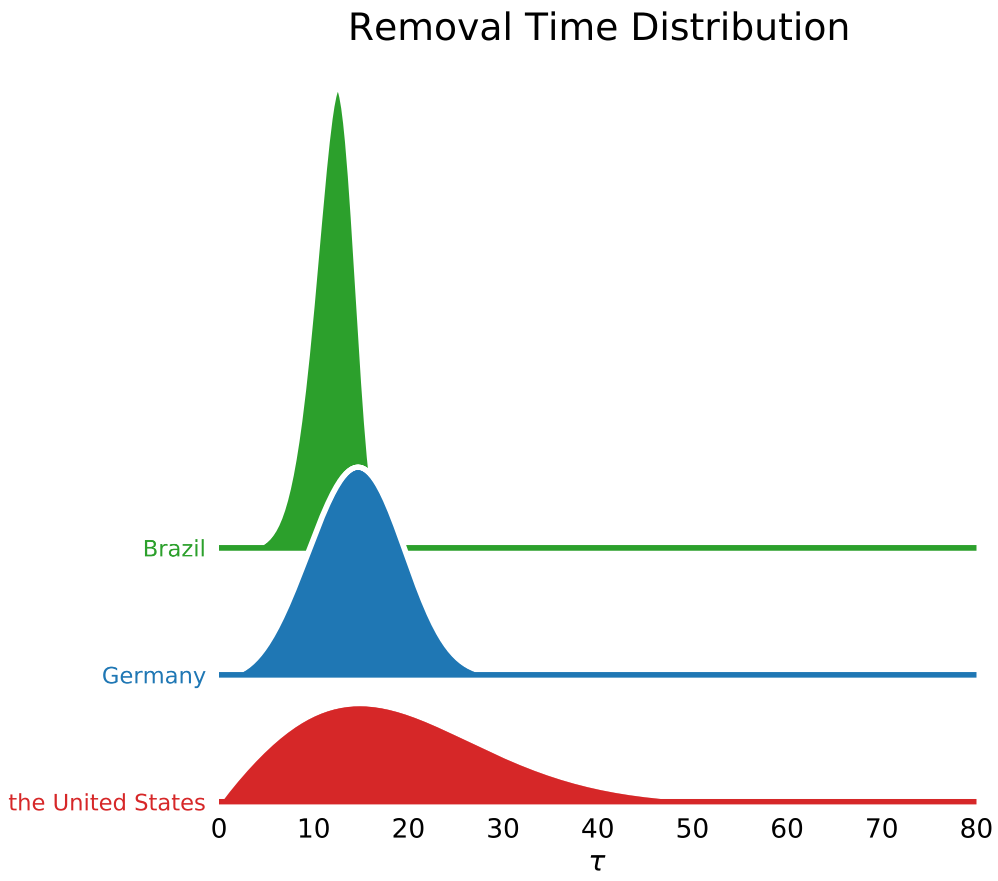

Supplementary Fig. 4: **Removal time distributions $\psi_{\text{rem}}(\tau)$ of the United States, Germany and Brazil.**

#### Section 1.4: Calibration of $k$

The disease transmission depends on the contact matrix adjustment parameter $k$ which is related to the government intervention policy and the public response, but there is no survey data to show how $k$ is affected by these factors. To solve this issue, we calculate the values of $k$ on each day before onset of vaccine immunity (the 14 days after the onset of vaccination, because 14 days is the average time before our bodies build effective immunity after the first dose of vaccination) by taking real data from Ref. [6] into Eqs. (1–2) in main text. And the time series of $k$ are presented (Supplementary Fig. 5, blue and red dots).

No data shows the detailed vaccine allocations of different age groups, so we cannot calculate and predict the values of $k$ after onset of vaccine immunity. For simplicity, we hypothesize that $k$ maintains a constant value with a 90% confidence interval during this period. To find the constant $k$ and corresponding confidence interval, we consider finding a consecutive time series of $k$ whose average value can lead to the calculation of the first 14 days after vaccination onset very close to the real data. But the mean value of the $k$ time series during the first 14 days of vaccination cannot achieve a satisfying result, so we decide to expand the $k$ series by testing all the dates before the onset of vaccination and selecting the date, from which to the onset of vaccine immunity the average $k$ can lead the calculation of the first 14 days after vaccination onset closest to the real data, as the starting date (Supplementary Fig. 5 red dots). Then we can obtain this constant $k$ and its 90% confidence interval by averaging and bootstrapping the time series of $k$ from the starting date to the onset of vaccine immunity (Supplementary Fig. 5 red lines and the intervals). The detailed onset of vaccination, onset of vaccine immunity and the chosen starting date of different countries are presented (Supplementary Table 2). Meanwhile, the calculated constant $k$ and the corresponding confidence intervals of different countries can also be demonstrated (Supplementary Fig. 5).

| Country | Onset of Vaccination | Onset of Vaccine Immunity | Chosen Starting Date |
|---|---|---|---|
| the United States | 20 December 2020 | 3 January 2021 | 17 December 2020 |
| Germany | 27 December 2020 | 10 January 2021 | 14 December 2020 |
| Brazil | 17 January 2021 | 31 January 2021 | 14 January 2021 |

Supplementary Table 2: **List of onset of vaccination [10], onset of vaccine immunity and the chosen starting date of the United States, Germany and Brazil.**

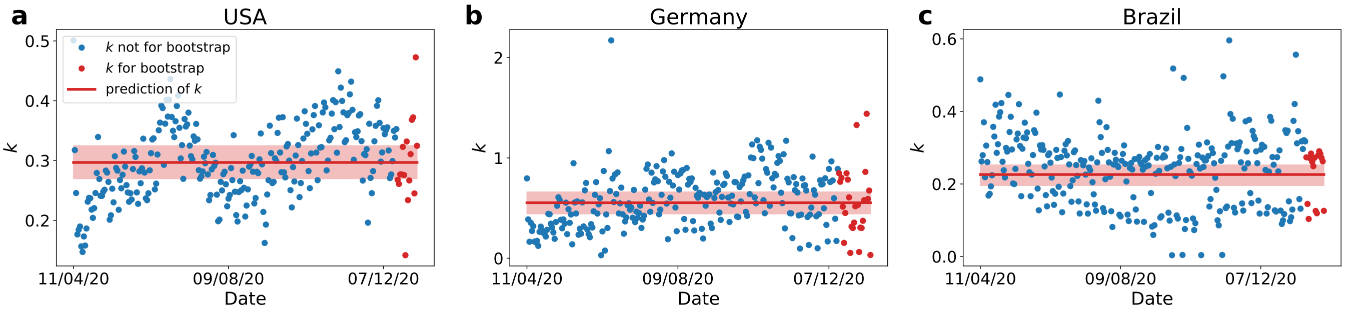

Supplementary Fig. 5: **Calculation and predictions of $k$.** The plots (blue and red) represent the calculated values of $k$ before the first immunity protection, the red plots represent the $k$ values chosen for bootstrap, the red lines and the corresponding intervals denote the predictions with 90% confidence levels for the constant prediction after the first immunity protection. **a** the United States; **b** Germany; **d** Brazil.

#### Section 1.5: $\sigma$ and $\eta$

Typically, after two weeks of full vaccination, the human body will build immunity against SARS-CoV-2, and some protection will appear 14 days after the first dose of vaccine [11]. For instance, Pfizer's two-dose vaccines are 95% effective in preventing symptomatic COVID-19 infection; however, studies have shown that the first dose is only 52% effective [12]. In our model, the delay from S to V after vaccination should satisfy a time distribution, but this distribution cannot be reflected by any currently available data source; therefore, we consider choosing a constant (an average delay) to characterize this process. There are two types of selection for consideration: 14 days after the full vaccination (95% of vaccinated people can be protected against the virus) and 14 days after the first dose (52% of vaccinated people acquire immunity) [11;12]. Because the time delay when approximately half of the vaccinated individuals gain immunity (the latter selection) has higher probability to be close to the average delay to build strong immunity protection, we choose 14 days after the first dose as the average time delay $\sigma$ for the vaccination process and 95% as the vaccine efficacy $\eta$.

#### Section 1.6: Calibration of rollout speed

We determined the fixed rollout speed as the average vaccination rate between the vaccination onset date (United States: 20 December 2020; Germany: 27 December 2020; Brazil: 17 January 2021) and 24 April 2021, from which we can obtain the rollout speed of different countries in our model (United States: 0.334%; Germany: 0.193%; Brazil: 0.129%) [10].

#### Section 1.7: Infection Fatality Rate and Calibration of $\mu$

As it is difficult to directly collect IFR data for current survey projects, the current data sources mainly present the case fatality rate (CFR, i.e., number of deaths divided by the number of confirmed cases). Therefore, we hypothesize that the distribution of the CFR is proportional to that of the IFR with a coefficient $\epsilon$. Meanwhile, the epidemic data of only a few countries included death proportions in different age groups (others only showed the death numbers but not the proportion); therefore, a linear proportional relationship is assumed to exist between the IFR distribution of one country and that of Spain with a coefficient $\varepsilon$ (data are only chosen from the CFR distribution of Spain, and we defined the CFR distribution in Spain as unit IFR distribution $\theta$, and actual IFR distribution in Spain equals to $\theta$ multiplied by the coefficient $\epsilon$) [13;14]. Then, the IFR at time $t$ can be expressed as $\epsilon\varepsilon\theta_l$ and we can obtain:

$$d_l(t) = \int_{-\infty}^{t} \mu\theta l d r_l(t'), \tag{S11}$$

where $d_l(t)$ is the proportion of death in the age group $l$, and $\mu = \epsilon\varepsilon$. As for the unit IFR distribution, the age division in real data is different from our model, then we interpolate them by linear method, and finally obtain IFR of each age group (Supplementary Fig. 6). Furthermore, note that there exists a little peak around the 10–14-year age group, which can affect the years of life lost discussed below, according to the data (Supplementary Fig. 6) [14].

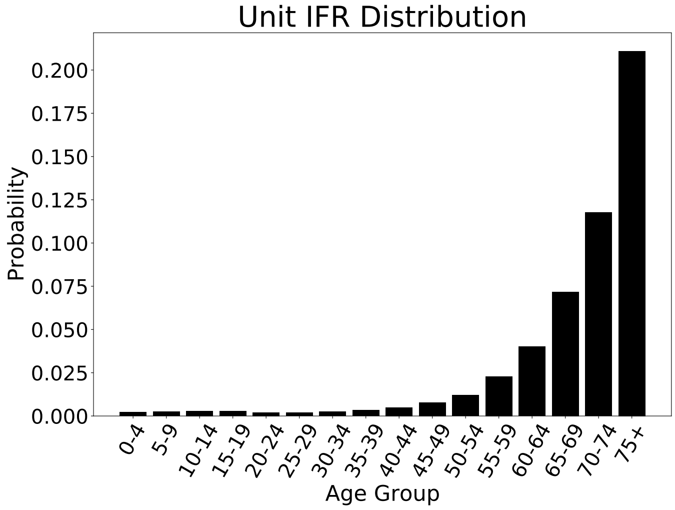

Supplementary Fig. 6: **Unit IFR distribution.** Overall, unit IFR increases with age, however, there still exists a little peak around the 10–14-year age group.

We put real death data into Eq. S11 and calculate the values of $\mu$ during 60 days before the onset of immunity, then obtain the mean value and its 90% confidence interval by bootstrapping method (Supplementary Fig. 7).

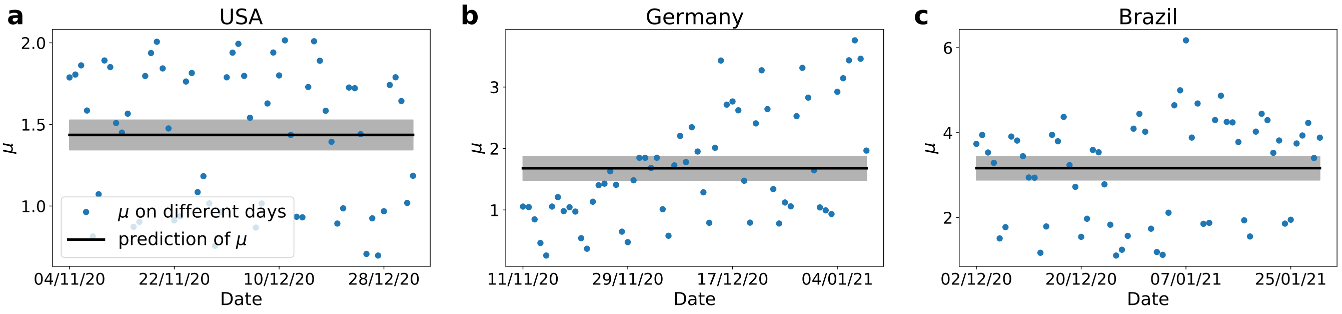

Supplementary Fig. 7: **The calculation and prediction of $\mu$ of the three countries:** The blue plots represent the calculated values of $\mu$ before the first immunity protection, the black lines and the corresponding intervals denote the predictions with 90% confidence levels for the constant prediction after the first immunity protection. **a** the United States; **b** Germany; **d** Brazil.

#### Section 1.8: Life Expectancy

To calculate years of life lost (YLL), the life expectancy of all the three countries are required (Supplementary Table. 3) [15]. YLL is defined as the time length from an individual's death to the life expectancy of his/her country. In our model, if the individual, whose age is beyond the life expectancy of his/her country, passes away, YLL will be set to 0.

| Country | Life Expectancy |
|---|---|
| the United States | 78.5 |
| Germany | 81.7 |
| Brazil | 75.9 |

Supplementary Table 3: **List of life expectancy of the United States, Germany and Brazil.**

#### Section 2: Steady-state Prediction

The whole spreading dynamics is considered under the non-Markovian framework, in which the future states are determined by both of the current and previous states. Furthermore, the introduction of vaccines which can reduce the susceptible population will further prevent the transmission of COVID-19 disease. Therefore, at a certain time point, there exists a functional relation between the vaccine allocation and the prediction of steady state, and the corresponding partial derivative of steady-state prediction with respect to vaccine allocation can be also calculated.

If the population get vaccinated with a vaccine distribution $\varpi$ (whose $l$-th element represents the proportion of the individuals of age group $l$ who got vaccinated with vaccine amount $\vartheta dt$ during an infinitesimal time interval $[t-\sigma, t-\sigma+dt)$) at time $t-\sigma$, and the bodies of vaccinated individuals will build protection against the virus at $t$ on average, then the vector $\tilde{c}$ which can characterize the steady state could satisfy the following self-consistent equations,

$$\tilde{c}_l = q_l^* - \varphi_l - (s_l^* - \varphi_l)\exp[-A_l(\lambda\tilde{c} - \lambda r^* - \lambda i^* + \lambda^* \circ i^*)], \tag{S12}$$

$$\varphi_l = \frac{\eta\vartheta\varpi_l e^{-\int_{t-\sigma}^{t} j_l(t')dt'}}{p_l}dt, \tag{S13}$$

$$A = kH \circ p, \tag{S14}$$

where $\tilde{c}$ is a vector whose $l$th element denotes the cumulative infection proportion in age group $l$ at steady state, $\tilde{c}_l$ is the $l$th element of vector $\tilde{c}$, $s_l^*$ is the susceptible proportion of age group $l$ at time $t$, $r^*$ and $i^*$ are two vectors whose elements denotes the removed and infected proportion of the corresponding age group at time $t$, respectively, $q_l^*$ means the proportion of who have not been vaccinated in the age group $l$ until time $t$, $A_l$ is the vector on the $l$th row of matrix $A$ which integrates constant $k$, contact matrix $H$ and age distribution $p$, $\varphi_l$ denotes the proportion of individuals, who get immune during the time interval $[t-\sigma, t-\sigma+dt)$, in susceptible population, $\eta$ is the efficacy of vaccines, "$\circ$" is the symbol of Hadamard product, $\lambda$ is the effective infection rate for the future infected individuals who will be infected after time $t$, $\lambda^*$ is a vector whose $l$th element denotes the average effective infection rate of the current infected people in age group $l$, $\lambda$ and $\lambda^*$ can be calculated by

$$\lambda = \int_0^{+\infty} \omega_{\text{inf}}(\tau)\Psi_{\text{rem}}(\tau)d\tau, \tag{S15}$$

$$\lambda^* = \int_0^{+\infty}\int_\tau^{+\infty} c'(t-\tau)\omega_{\text{inf}}(\tau')\Psi_{\text{rem}}(\tau')d\tau' d\tau \oslash i^*. \tag{S16}$$

where "$\oslash$" is the symbol of Hadamard division.

Then the predictions of FCI, FCD and FYLL can also be obtained as follows:

$$\chi_{\text{FCI}} = \tilde{c}^T p, \tag{S17}$$

$$\chi^*_{\text{FCD}} = \mu\tilde{c}^T(p \circ \theta), \tag{S18}$$

$$\chi^*_{\text{FYLL}} = \mu\tilde{c}^T(p \circ \theta \circ \phi), \tag{S19}$$

where $\chi_{\text{FCI}}$ is the prediction of FCI, $\chi^*_{\text{FCD}}$ and $\chi^*_{\text{FYLL}}$ are the predictions of FCD and FYLL, respectively, $\theta$ and $\phi$ are unit IFR distribution and YLL distribution in different age groups, respectively (Supplementary Fig. 6). Therefore, the steady state of the non-Markovian age-stratified SIR model can be predicted accurately by any transient state with its current and previous processes.

Moreover, the algorithm to search the parameters which can lead to the minimum results also need the corresponding partial derivative which can be expressed as

$$\frac{\partial \tilde{c}_l}{\partial \varphi_m} = [\lambda A_l \frac{\partial \tilde{c}}{\partial \varphi_m}(s^*_l - \varphi_l) + \delta_{lm}] \exp[-A_l(\lambda\tilde{c} - \lambda r^* - \lambda i^* + \lambda^* \circ i^*)] - \delta_{lm}. \tag{S20}$$

where $\delta_{lm}$ is Kronecker delta function which satisfies

$$\delta_{lm} = \begin{cases} 1 & l = m \\ 0 & l \neq m \end{cases} \tag{S21}$$

To verify the ability of steady-state prediction, we calculate the non-Markovian processes with a constant value of $k$. And we predict the three final metrics, FCI, FCD and FYLL, according to the current states along with the corresponding previous information, and all the predictions can be consistent with the steady state from the full simulation of the non-Markovian model, meaning that the predictions by our method are completely accurate (Supplementary Fig. 8).

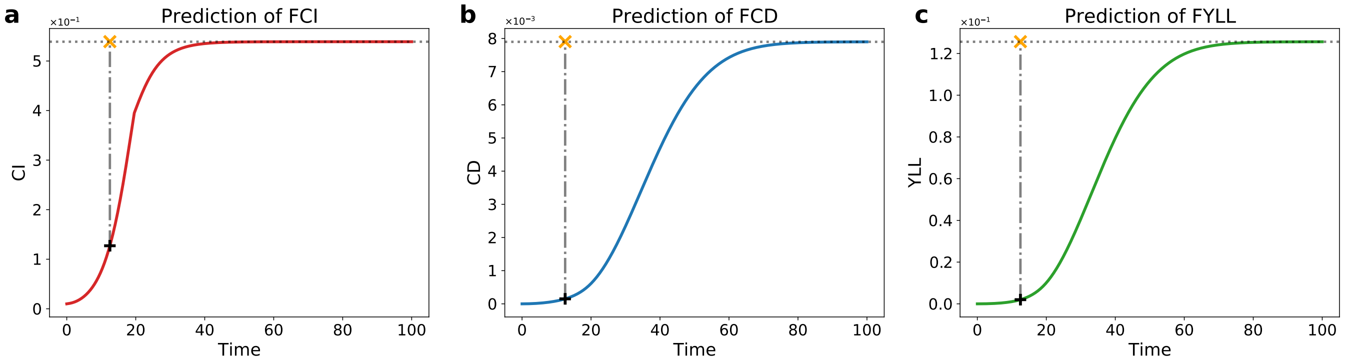

Supplementary Fig. 8: **Verification of the ability to steady-state prediction.** **a**–**c**, predictions of FCI, FCD and FYLL with one single vaccination at a certain time point. The markers " + " point the time when the steady states are predicted, and markers " × " indicate the corresponding steady states predicted. The nonsmooth point after vaccination in **a** reflects the protection built by vaccines after certain days, and the main reason of the absence of unsmooth point in **b** and **c** is due to the convolution calculation to obtain cumulative-death and YLL.

### Section 3: Steady-state Optimization

The detailed optimization algorithm for the steady-state prediction in our work is Sequential Quadratic Programming[7;16]. According to Eqs. S12–S21, there must exists three functions which can describe the relationship between the predictions of FCI, FCD and FYLL as follows:

$$\chi_{\mathrm{FCI}} = f_{\mathrm{FCI}}(\varphi), \tag{S22}$$

$$\chi_{\mathrm{FCD}} = f_{\mathrm{FCD}}(\varphi), \tag{S23}$$

$$\chi_{\mathrm{FCYLL}} = f_{\mathrm{FYLL}}(\varphi), \tag{S24}$$

where the $l$-th element of vecotr $\varpi$ denotes the proportion of vaccinated individuals in susceptible population.

If the vaccine supply is limit to a amount $\vartheta dt$, the constraint condition could be

$$0 \leq \varphi^T p \leq \eta\vartheta e^{-\int_{t-\sigma}^{t} j_l(t')dt'} dt, \tag{S25}$$

Note that, $\chi_{\mathrm{FCI}}$, $\chi_{\mathrm{FCD}}$ and $\chi_{\mathrm{FYLL}}$ are just the predictions for FCI, FCD and FYLL on the condition that there are no more vaccine for the further vaccination and $k$ remains constant after the current vaccination.

We choose optimize $\chi_{\mathrm{FCI}}$ as an example, and according to Sequential Quadratic Programming, the target of optimization is as follows:

$$\begin{aligned} &\min_{\varphi} f_{\mathrm{FCI}}(\varphi) \\ \text{subject to } & b(\varphi) \geq 0 \\ & c(\varphi) \geq 0 \end{aligned} \tag{S26}$$

where $b(\varphi) = \varphi^T p$ and $c(\varphi) = \eta\vartheta e^{-\int_{t}^{t+\sigma} j_l(t')dt'} dt - \varphi^T p$, The Lagrangian for this problem is described as

$$L(\varphi, x, y) = f_{\mathrm{FCI}}(\varphi) - xb(\varphi) - yc(\varphi) \tag{S27}$$

where $x$ and $y$ are Lagrange multipliers for this problem. At an iterate $\varphi_l$, an appropriate search direction $g_l$ is defined as a solution to the quadratic programming subproblem

$$\begin{aligned} \min_{g} \; & f_{\mathrm{FCI}}(\varphi_k) + \nabla f(\varphi_k)^T d + \frac{1}{2} g^T \nabla^2_{ll} \mathcal{L}(\varphi_{ml}, x_l, y_l) g \\ \text{s.t. } & b(\varphi_l) + \nabla b(\varphi_l)^T g \geq 0 \\ & c(\varphi_l) + \nabla c(\varphi_l)^T g \geq 0 \end{aligned} \tag{S28}$$

Please note that the term $f_{\mathrm{FCI}}(\varphi_l)$ in the equation above may be left out for the minimization problem, because it is constant under the $\min_{g}$ operator[16]. After finding the optimal $\varphi$, the corresponding vaccine distribution vector $\varpi$ can be calculated by Eq. S13.

#### Section 4: Pandemics in Other Countries

We also show the simulation results of Germany, and Brazil (mean rollout speed 0.193% for Germany, and 0.129% for Brazil, which are calculated from their vaccination onset day to 24 April 2021 [10]).

#### Section 4.1: Pandemics in Germany

As for the prevention of SARS-COV-2 virus spreading in Germany, OVS can also perform the best (Supplementary Fig. 9). For the detailed vaccine allocation of OVS, the 15–19-year age group should be vaccinated first followed by the 40–44-year age group several days later. After other age groups begin to get vaccinated, the vaccination of the 15–19-year age group should remain at a relatively low level for a long period. For minimizing FCI, 30–34-, 35–39-, 20–24- and 45–49-year age groups should be vaccinated in order. For minimizing FCD, 30–34-, 35–39-, and 50–54-year age groups need to be vaccinated after a period of vaccination of the 40–44-year age group. For minimizing FYLL, 30–34-, 35–39-, 45–49- and 50–54-year age group should be vaccinated (Supplementary Fig. 10).

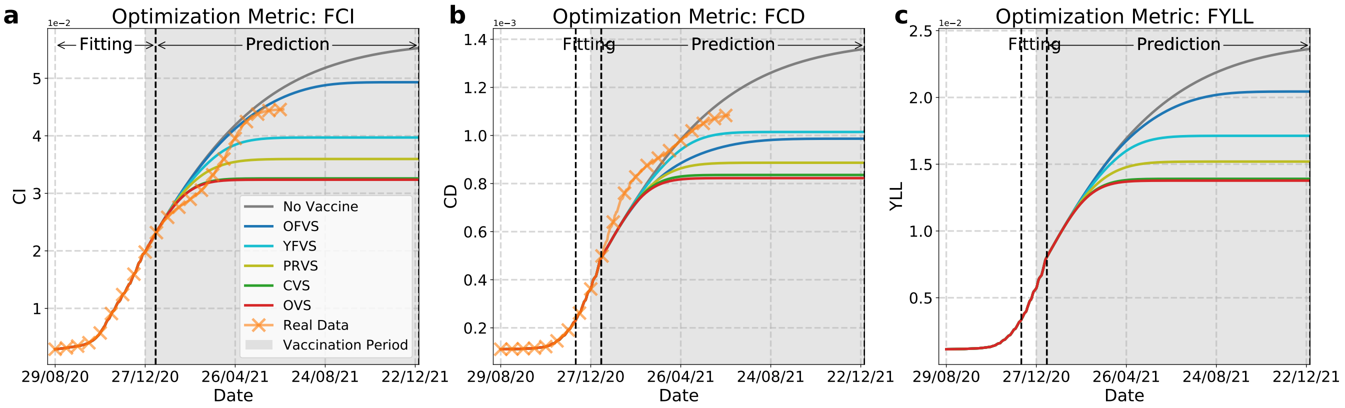

Supplementary Fig. 9: **Predictions of COVID-19 spread in Germany under different vaccination strategies.** Panels **a**–**c**, Growth curves of cumulative-infection (CI), cumulative-death (CD), and years of life lost (YLL).

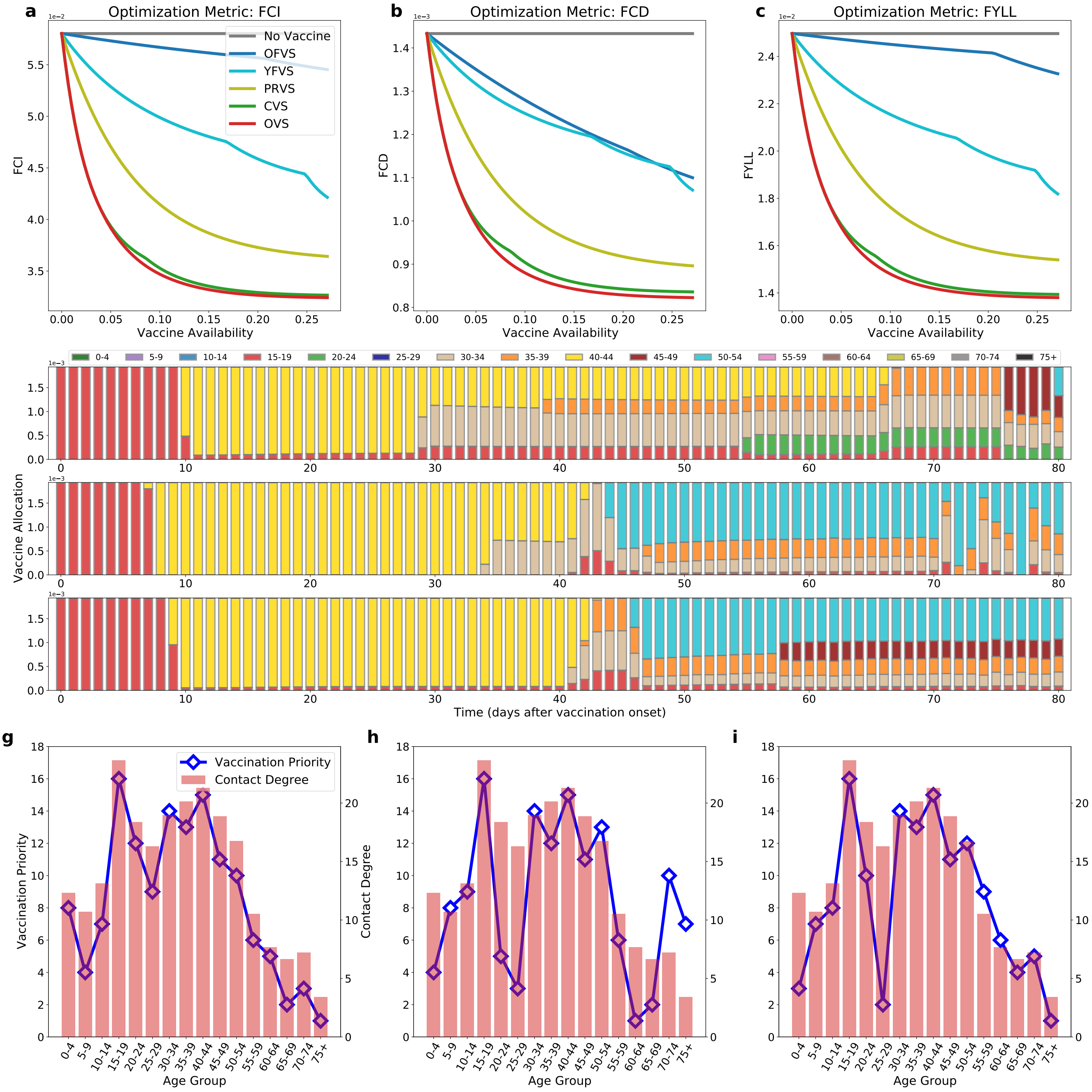

Supplementary Fig. 10: **Strategy comparison, vaccine distribution of the optimal vaccination strategy (OVS), and vaccination priorities for age groups with respect to vaccine availability in Germany.** (**a**–**c**) The final cumulative-infection (FCI), final cumulative-death (FCD), and final years of life lost (FYLL) under different vaccine availabilities. (**d**–**f**) Vaccine distribution in different age groups in the OVS with different metrics, FCI, FCD, and FYLL, when the vaccine is available up to given days (horizontal axis). The bar on each single day in the subfigures represents the vaccine distribution to the corresponding age group. (**g**–**i**) Vaccination priorities for the age groups obtained through the vaccine distributions shown in (**d**–**f**), with different targets (FCI, FCD, and FYLL), compared with the contact degree of the age groups in Germany.

### Section 4.2: Pandemics in Brazil

There exist some deviation of the calculation of Brazil from the real data, this can reflect that the value of $k$ in Brazil remains a very high level, and this may be mainly because the value of $k$ in reality increased after the vaccination. As for the prevention of SARS-COV-2 virus spreading in Brazil, OVS can also perform best (Supplementary Fig. 11). For the detailed vaccine allocation of OVS in Brazil, the 15–19-year age group should be vaccinated first and then around the 20th day of the vaccination, the 10–14-year age group need to start vaccination together with the 15–19-

year age group. Then the vaccination should be followed by the 25–29- and 20–24-year age group for minimizing FCI, the 25–29- and 75+-year age group for minimizing FCD and the 25–29-, 20–24- and 5–9-year age group for minimizing FYLL (Supplementary Fig. 12).

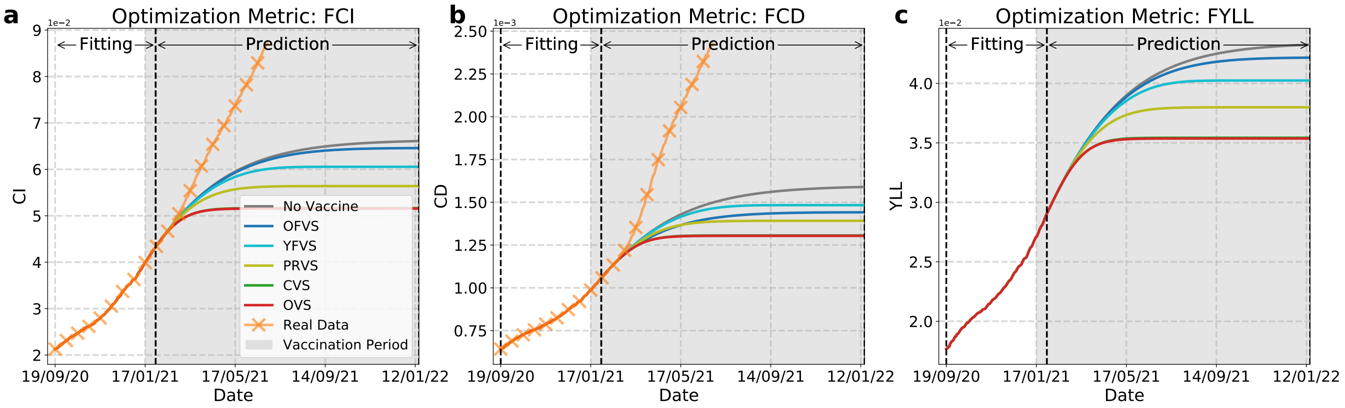

Supplementary Fig. 11: **Predictions of COVID-19 spread in Brazil under different vaccination strategies.** Panels **a**–**c**, Growth curves of cumulative-infection (CI), cumulative-death (CD), and years of life lost (YLL).

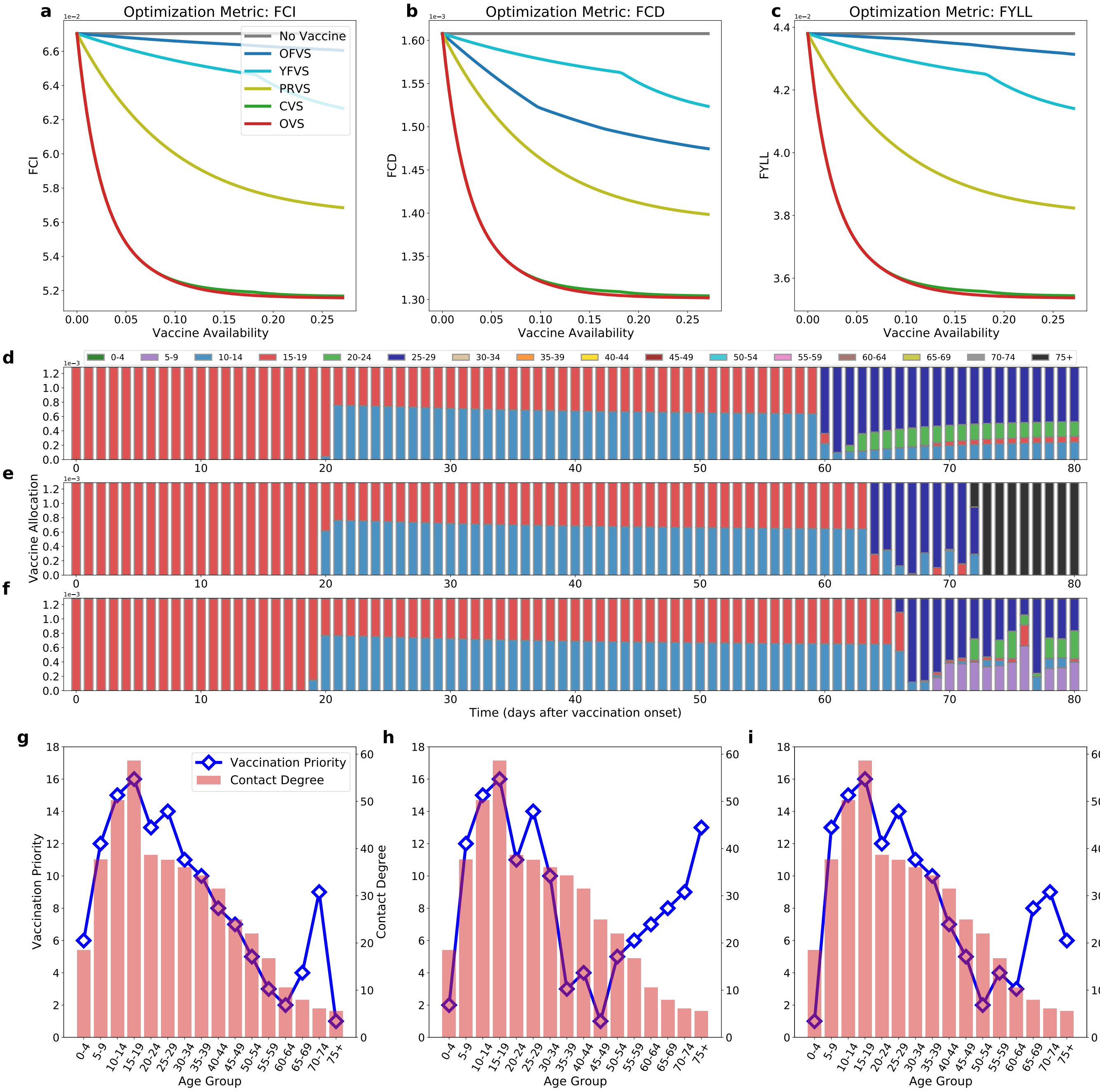

Supplementary Fig. 12: **Strategy comparison, vaccine distribution of the optimal vaccination strategy (OVS), and vaccination priorities for age groups with respect to vaccine availability in Brazil.** (**a**–**c**) The final cumulative-infection (FCI), final cumulative-death (FCD), and final years of life lost (FYLL) under different vaccine availabilities. (**d**–**f**) Vaccine distribution in different age groups in the OVS with different metrics, FCI, FCD, and FYLL, when the vaccine is available up to given days (horizontal axis). The bar on each single day in the subfigures represents the vaccine distribution to the corresponding age group. (**g**–**i**) Vaccination priorities for the age groups obtained through the vaccine distributions shown in (**d**–**f**), with different targets (FCI, FCD, and FYLL), compared with the contact degree of the age groups in Brazil.

### Section 5: Vaccination Duration with Different Efficacies

The efficacy of vaccines can influence the control of epidemic prevention, for example, the duration of vaccination. We define effective vaccination duration (EVD) as the time length from the vaccination onset during which the vaccination can reduce the number of new CI, CD or YLL to be less than 1% of that at the onset of vaccination. Higher efficacy can lead to faster decrease of new CI, CD and YLL by using the vaccination distributions generated by OVS (Supplementary Fig. 13a–c), resulting in lower EVD with approximate linear relationships between them (Supplementary Fig. 13a–c, insets). In addition, reducing new CI to a relatively low level (93 days for 50% efficacy, 62 days for 100% efficacy) usually takes a month less than reducing new CD (123 days for 50% efficacy, 94 days for 100% efficacy). As for reducing the new YLL (120 days for 50% efficacy, 90 days for 100% efficacy), it just takes around 3 days less than reducing the new CD. Meanwhile, we can quantify the effectiveness of vaccination as the proportion of reduced FCI, FCD and FYLL under vaccination with respect to FCI, FCD and FYLL without vaccination, respectively. The effectiveness increases linearly with the efficacy, and can at least reduce 20% of FCI, FCD and FYLL by OVS with efficacy above 50% (Supplementary Fig. 13a–c, insets).

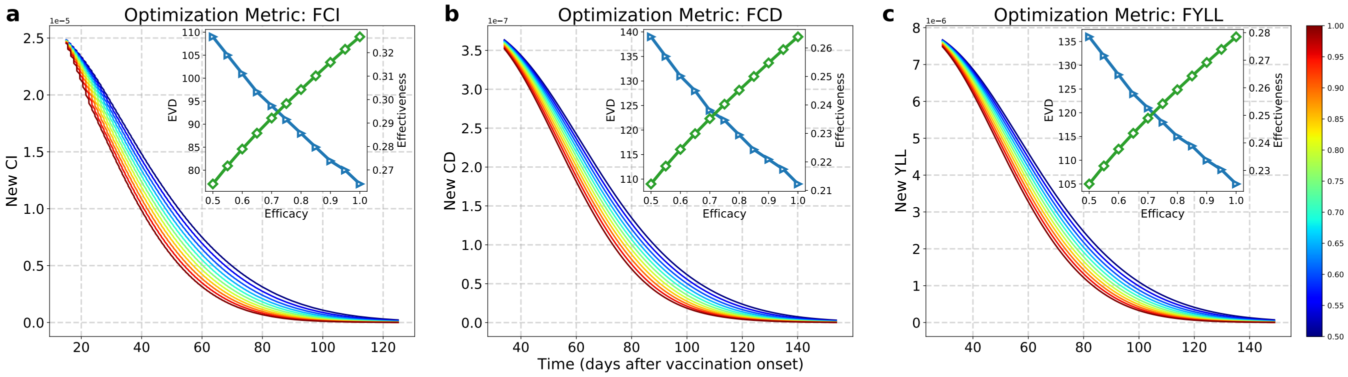

Supplementary Fig. 13: **Effect of vaccine efficacies under OVS. a–c** Curves of new CI, CD and YLL under different efficacies (from 50% to 100% with the increment of 5%, from dark blue to dark red color). Insets: EVD (the blue lines with triangle markers) and effectiveness (the green lines with diamond markers) with respect to efficacy.

### Section 6: Optimal Vaccination Effects in Different Countries

We also calculate the effectiveness of vaccines on prevention in three different countries, the United States, Germany and Brazil with the five strategies, OVS, CVS, PRVS, OFVS and YFVS. According to the real data, we set the vaccination rollout rate of the United States, Germany and Brazil as 0.334%, 0.193% and 0.129% of their own total population per day and then set the vaccination starting on as 20 December 2020, 27 December 2020 and 17 January 2021, and ending one year later, on 19 December 2021, 26 December 2021 and 16 January 2022, respectively [10]. The radar charts present the influence of different strategies in different countries for the three prevention objectives (Supplementary Fig. 14a–c). In all the three countries, OVS can lead to the best effectiveness. The areas of the curves can reflect the overall effectiveness of vaccinations, including every vaccination strategy, on the prevention of a given country. The vaccination has the largest effect on the pandemic control of Germany and the smallest influence on that of Brazil. Meanwhile, the total effectiveness in the three countries has a significant enhancement for reducing FCD by OFVS in contrast to those for reducing FCD by other methods.

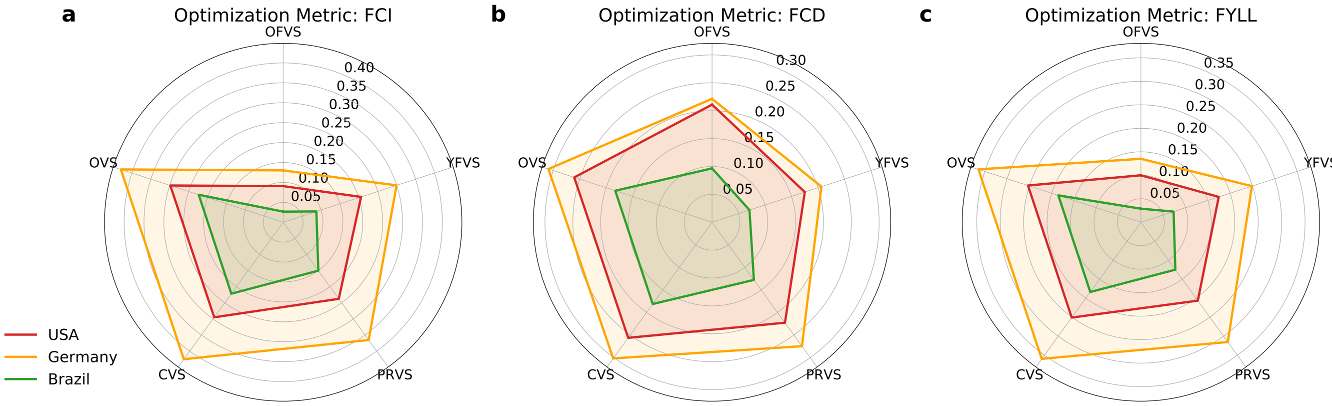

Supplementary Fig. 14: **Comparison of vaccination effectiveness in different countries. a–c** Radar chart showing the effectiveness under five vaccination strategies for reducing FCI, FCD and FYLL for the United States, Germany and Brazil.

#### Section 7: PEV for Reducing FCI and FYLL

We can also obtain the results of PEV for reducing FCI and FYLL (Supplementary Figs. 15–16). The results for reducing the two metrics have the same phenomena as that for reducing FCD in the main text, enable us to obtain the same conclusions, and enhance the feasibility of our approaches.

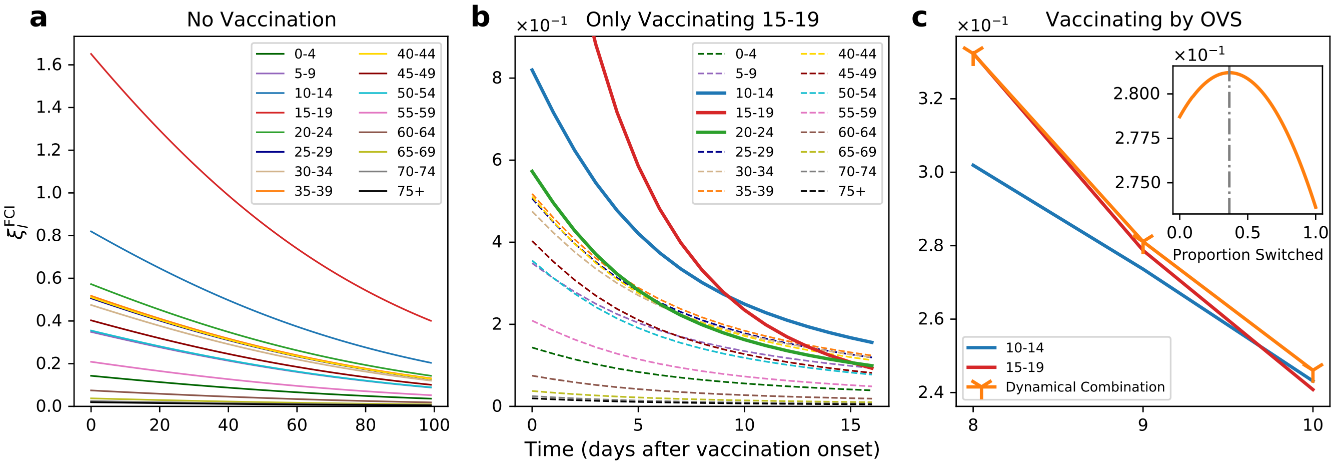

Supplementary Fig. 15: **Mechanisms of the optimal vaccination strategy (OVS) for reducing the final cumulative deaths (FCI).** (**a**) Synchronous change of prevention efficiency with vaccine (PEV) for reducing the FCI of each age group with the evolution of spreading dynamics when vaccination is not implemented. (**b**) Dynamic changes in the PEV of each age group while the 15–19-year age group is vaccinated. Note that the PEV of 15–19-year age group declines fastest. (**c**) Detailed dynamic changes in the PEVs of the 10–14- and 15–19-year age groups, and their combination. Inset: orange solid line shows the changes in the PEV for reducing the FCI under different vaccine percentages switched from the 15–19-year age group to the 10–14-year age group on the 9th day (0 indicates that all vaccines are provided for the 15–19-year age group, and 1 indicates that all vaccines are supplied to the 10–14-year age group). The vertical grey-dashed line passing the maximum represents the percentage generated by the OVS as shown in Fig. 3d in the main text.

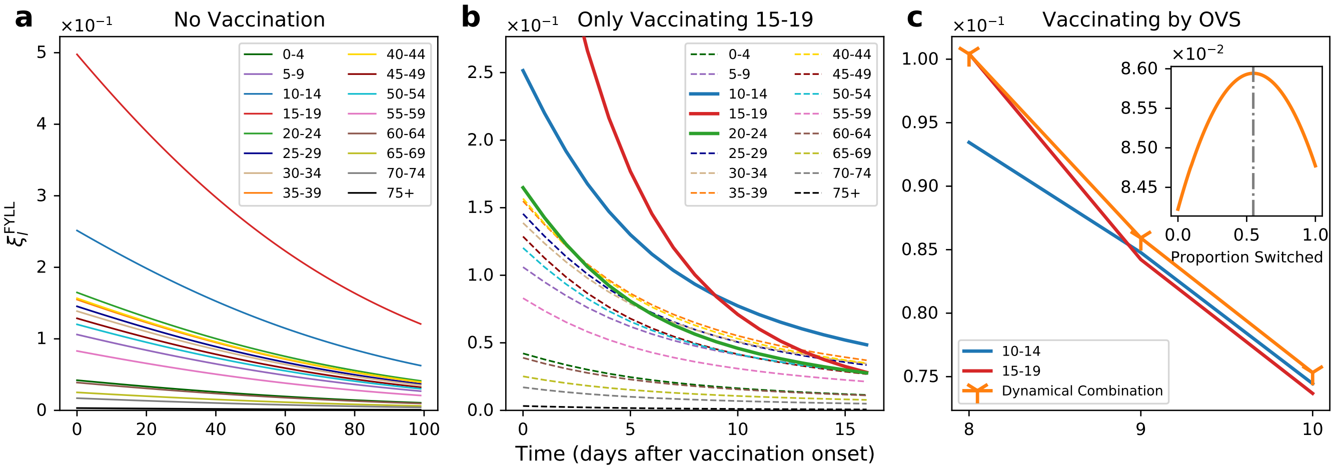

Supplementary Fig. 16: **Mechanisms of the optimal vaccination strategy (OVS) for reducing the final cumulative deaths (FYLL).** (**a**) Synchronous change of prevention efficiency with vaccine (PEV) for reducing the FYLL of each age group with the evolution of spreading dynamics when vaccination is not implemented. (**b**) Dynamic changes in the PEV of each age group while the 15–19-year age group is vaccinated. Note that the PEV of 15–19-year age group declines fastest. (**c**) Detailed dynamic changes in the PEVs of the 10–14- and 15–19-year age groups, and their combination. Inset: orange solid line shows the changes in the PEV for reducing the FYLL under different vaccine percentages switched from the 15–19-year age group to the 10–14-year age group on the 9th day (0 indicates that all vaccines are provided for the 15–19-year age group, and 1 indicates that all vaccines are supplied to the 10–14-year age group). The vertical grey-dashed line passing the maximum represents the percentage generated by the OVS as shown in Fig. 3f in the main text.
